# Automated Screening for Generalized Anxiety Disorder Using Voice Analysis and Machine Learning

**DOI:** 10.64898/2026.11.18.26367837

**Authors:** Arman Khoshnevis, Mohsen Zayernouri, Leapetswe Malete, Dale Joachim, Chelsi Ricketts, Reginald Ocansey, Rosemary Chizobam Muomah, Dawn Tladi, Joyce Sifa Ndabi, Maryam Naghibolhosseini

## Abstract

This study evaluated machine learning models for screening elevated generalized anxiety symptoms from remotely collected neutral read speech in 1,788 adults from Botswana, Ghana, Nigeria, and Tanzania. Elevated symptoms were defined by Generalized Anxiety Disorder 7-item (GAD-7) questionnaire scores ≥5. Nine classifiers were assessed across nine scenarios defined by speech-task configuration (short sentence, long passage, or both tasks) and sex stratification under participant-separated, repeated nested cross-validation (CV). Acoustic-only models used the extended Geneva Minimalistic Acoustic Parameter Set, and context-augmented models added age, self-reported health, and sex where applicable. With feature selection, the highest mean area under the receiver operating characteristic curve (AUC-ROC) was 0.594 for acoustic-only models and 0.699 for contextaugmented models, the latter in the female-only combined-task scenario. Shapley additive explanations (SHAP) identified fundamental frequency descriptors as prominent acoustic predictors, while health status and age were the leading predictors in context-augmented models. Performance varied across scenarios, and feature selection provided no consistent benefit. Secondary regression poorly estimated continuous symptom severity. These findings indicate limited discrimination from neutral speech acoustics alone and improved performance with participant context, warranting external validation.

## I. INTRODUCTION

**G**eneralized Anxiety Disorder (GAD) is a pervasive mental health condition that imposes a substantial global socioeconomic burden and affects individual quality of life [1], [2], [3]. Anxiety disorders affect approximately 4.7% of the global population, making them the most prevalent category of mental disorders worldwide [4]. GAD onset often peaks during the transition to young adulthood, when early intervention can improve long-term outcomes [5], [6], [7]. However, conventional clinical assessment relies on clinicianled interviews and self-report measures that are susceptible to recall bias, clinician subjectivity, and social desirability effects [8], [9]. These limitations motivate objective, scalable, and non-invasive screening tools suitable for naturalistic and remote settings [10], [11], [12].

These screening needs have prompted growing interest in digital biomarkers for mental health assessment [13], [14], [15]. Among these, vocal biomarkers offer a promising approach grounded in the psychophysiological links between emotional states and speech production [16]. Anxiety-related increases in muscle tension and changes in respiratory patterns can alter vocal production and acoustic features [17], [18], [19]. However, associations between these features and anxiety presence or severity remain inconsistent. This heterogeneity may reflect differences in the anxiety construct, speech task and elicitation conditions (including stress-elicited, neutral, or spontaneous speech), sample size, and participant demographic or health characteristics [20], [21], [22], [23], [24], [25].

To characterize complex patterns across high-dimensional acoustic features, research has increasingly adopted machine learning (ML) and deep learning (DL) approaches within affective computing [26], [16]. Applications include screening of generalized anxiety [27], social anxiety [28], [29], and other anxiety-related disorders [17], [30]. Beyond anxiety, these approaches address depression [31], [32], [33], post-traumatic stress disorder [34], [35], and other psychiatric and neuropsychiatric outcomes [36], [37], alongside general emotion recognition [38], [39], [40], [41]. These studies draw on clinical interview corpora such as DAIC-WOZ [42], crowd-sourced datasets such as DEPAC [43], emotion-recognition collections such as EmoDB [44], and study-specific recordings from demographically diverse populations [45], [46], [47].

Across these applications, feature representations range from handcrafted acoustic descriptors to learned embeddings and multimodal combinations. Handcrafted acoustic low-level descriptors (LLDs) capture prosodic, energy, spectral/cepstral, and glottal properties [47], [39], [11], often organized into established feature sets such as Geneva minimalistic acoustic parameter set (GeMAPS) and its extended version (eGeMAPS) [48], ComParE [49], and emobase [50]. Neural approaches offer learned representations, including Wav2Vec2 embeddings [51] and X-vectors [52], that complement conventional acoustic descriptors. It is worth mentioning that combining acoustic representations with linguistic, physiological, or behavioral inputs, such as text, heart rate variability, and gait measures, can further improve classification [29], [37], [53].

Using these feature representations, conventional ML classifiers, including support vector machines (SVM), random forests (RF), k-nearest neighbors (KNN), multilayer perceptrons (MLP), logistic regression (LR), and Gaussian process (GP) models, have been widely applied to emotion recognition and anxiety screening. Although such pipelines often perform well on acted emotion corpora [38], [40], findings are more heterogeneous in anxiety and internalizing disorder screening. McGinnis et al. [11] achieved 80% accuracy in detecting childhood internalizing disorders using speech elicited immediately after an anxiety-inducing trigger during the Trier Social Stress Task adapted for children (TSST-C). Baird et al. [17] found that anxiety-related acoustic effects in sustained vowels were more detectable at higher Beck Anxiety Inventory (BAI) scores, particularly when participants spoke while smiling. For adult social anxiety, Alon-Ronen et al. [28] reported an area under the receiver operating characteristic curve (AUCROC) of 0.83 using a GP classifier, with performance varying by utterance type and higher accuracy in single-sex than combined-sex samples. In a broader GAD-screening context, Teferra et al. [27] reported more modest discrimination from impromptu speech, with AUC-ROC increasing from 0.57 using acoustic and linguistic features to 0.62 after adding demographic variables. Collectively, these findings suggest that conventional ML can identify informative vocal patterns, but its screening performance depends on the symptom construct, severity range, speech task, population characteristics, and evaluation setting [54].

Within conventional ML, boosted tree models such as XGBoost, LightGBM, and CatBoost provide practical tools for capturing nonlinear dependencies in high-dimensional acoustic and multimodal features. Park et al. [29] combined acoustic and physiological features from virtual-reality-based social anxiety sessions, with LightGBM achieving an AUC-ROC of 0.819 for trait anxiety. In a more naturalistic setting, Sousa et al. [46] analyzed smartphone-based conversational speech from pregnant women and reported an AUC-ROC of 0.71 for depression and anxiety screening using LightGBM with Melfrequency cepstral coefficients (MFCCs) and openSMILE-derived features, although sensitivity remained limited under class imbalance. Feature selection also influenced performance as Requardt et al. [55] found that a compact RF-selected subset of acoustic features outperformed larger feature sets for recognizing emotions of drivers, including anxiety, from in-vehicle speech. Together, these studies support ensemble methods as a practical approach to structured acoustic and multimodal modeling, while highlighting the importance of feature selection and evaluation under realistic recording conditions.

In parallel, DL frameworks, including convolutional neural networks (CNNs), long short-term memory networks (LSTMs), attention mechanisms, and transformer-derived speech representations, have been explored to learn higher-level vocal representations and reduce reliance on handcrafted features. Pretrained representations, such as Wav2Vec2 and X-vectors, can encode information beyond conventional acoustic descriptors. Brueckner et al. [56] found that pretrained speech representations outperformed MFCC-based baselines, although the best anxiety-detection performance remained moderate, with an unweighted average recall (UAR) of 0.60. These approaches remain constrained by limited clinical datasets, noisy self-report labels, class imbalance, domain shifts, and computational cost. More broadly, credible screening via ML and DL approaches requires leakage-free validation, appropriate feature selection, careful handling of speaker and demographic factors, training-fold resampling, and performance metrics suited to class imbalance [26], [54], [57]. Despite these advances and considerations, the extent to which neutral read speech collected remotely captures information relevant to screening elevated generalized anxiety symptoms remains uncertain. In particular, questions remain about how discrimination changes when participant context is included and how these patterns vary across speech tasks and sex-stratification scenarios under participant-separated evaluation. To address these questions, we evaluate conventional ML models for screening elevated generalized anxiety symptoms from remotely collected neutral read speech in a large multi-country African cohort. We compare acoustic-only eGeMAPS features with context-augmented features incorporating demo-graphic and health information across nine scenarios defined by speech-task configuration and sex stratification. Shapley additive explanations (SHAP) characterize the relative contributions of acoustic and contextual predictors. A common modeling pipeline with participant-separated, repeated nested cross-validation (CV) supports these comparisons while reducing data leakage and tuning-related optimism. As a secondary analysis, corresponding regression models assess whether the same predictors can estimate continuous Generalized Anxiety Disorder 7-item (GAD-7) scores. Sections II–V present the methods, results, discussion, and conclusions, respectively.

## II. MATERIALS AND METHODS

### A. Participants and Study Design

The present cross-sectional study analyzed data from a multi-country remote investigation conducted during the COVID-19 pandemic among adults in Botswana, Ghana, Nigeria, and Tanzania. In the broader study, a comprehensive set of behavioral, health-related, cognitive, mental health, demographic, and speech data were collected from participants through the SurveyLex platform (Sonde Health, Inc., Boston, MA) using their own smartphones, tablets, laptops, or desktop computers. The present analysis focused specifically on screening elevated generalized anxiety symptoms from remotely collected neutral read-speech recordings using machine learning models. This work extends our earlier analysis of the same cohort, which focused on fundamental-frequency-related measures, by evaluating whether a broader acoustic representation can support generalized anxiety classification under naturalistic remote recording conditions [20].

Study procedures were approved by the relevant institutional review boards or ethics committees at Michigan State University, the University of Nigeria, the University of Botswana, the University of Ghana, and the University of Dar es Salaam. Trained research assistants recruited participants through electronic channels, including email and social media platforms. Eligible participants were adults aged 18 years or older who provided informed consent before participation.

### B. Demographics and Clinical Measures

The demographic and health-related variables considered in this study included sex, age, and self-reported health status. These variables were used to characterize the analytic cohort and, where appropriate, as participant-level predictors in the machine learning analyses. Country was recorded as Botswana, Ghana, Nigeria, or Tanzania; sex was recorded as male or female; and age was analyzed as a continuous variable. Self-reported health status was binarized, with “very good” and “excellent” responses grouped as good health and “poor,” “fair,” and “good” responses grouped as poor health.

Anxiety symptoms were measured using GAD-7, a self-report questionnaire assessing anxiety-related symptoms over the preceding two weeks [58]. Each item is scored from 0 to 3, yielding a total score from 0 to 21, with higher scores indicating greater symptom severity. For supervised classification, participants with scores of 5 or higher were classified as anxious (having elevated generalized anxiety symptoms), whereas those with scores below 5 were classified as non-anxious (non-elevated). This cutoff was retained to preserve sensitivity to mild symptoms [59], informed by our previous analysis of this cohort [20], and reduce class imbalance. For the secondary regression analysis, the original GAD-7 total score was retained as a continuous outcome.

Table I summarizes the demographic and clinical characteristics of the final analytic cohort. Overall, 1,788 adults were included, with a near-balanced sex distribution. Across the cohort, 1,096 participants were classified as non-anxious and 692 as anxious, indicating moderate class imbalance. Anxiety-label distributions varied by country, with the highest proportion of elevated symptoms observed in Botswana and the lowest in Tanzania. Self-reported health status also varied across countries and sex groups; 1,066 participants were categorized as having good health and 722 as having poor health, with Botswana showing the highest proportion of poor-health responses. Age distributions differed across countries, with younger average ages in Ghana and Botswana and older average ages in Nigeria and Tanzania. These cohort-level differences supported the inclusion of participant-level variables in context-augmented models and motivated sex-stratified modeling scenarios.

**TABLE 1.** Cohort demographics, self-reported health, and anxiety status by country and sex.

| **Country** | **Variable** | **Male** | **Female** | **Both** |
|---|---|---|---|---|
| **Botswana** | Participants | 159 | 274 | 433 |
|  | Health (good/poor) | 71/88 | 100/174 | 171/262 |
|  | Anxiety (non-anxious/anxious) | 86/73 | 115/159 | 201/232 |
|  | Age (years) | 24.98 ± 6.43 | 26.17 ± 9.38 | 25.73 ± 8.43 |
| **Ghana** | Participants | 290 | 189 | 479 |
|  | Health (good/poor) | 196/94 | 97/92 | 293/186 |
|  | Anxiety (non-anxious/anxious) | 182/108 | 103/86 | 285/194 |
|  | Age (years) | 23.64 ± 5.17 | 23.13 ± 5.56 | 23.44 ± 5.33 |
| **Nigeria** | Participants | 275 | 297 | 572 |
|  | Health (good/poor) | 197/78 | 206/91 | 403/169 |
|  | Anxiety (non-anxious/anxious) | 197/78 | 195/102 | 392/180 |
|  | Age (years) | 29.40 ± 9.65 | 28.95 ± 9.96 | 29.17 ± 9.81 |
| **Tanzania** | Participants | 194 | 110 | 304 |
|  | Health (good/poor) | 139/55 | 60/50 | 199/105 |
|  | Anxiety (non-anxious/anxious) | 144/50 | 74/36 | 218/86 |
|  | Age (years) | 30.66 ± 8.06 | 28.05 ± 7.57 | 29.72 ± 7.97 |
| **Overall** | Participants | 918 (51%) | 870 (49%) | 1788 |
|  | Health (good/poor) | 603/315 | 463/407 | 1066 (60%)/722 (40%) |
|  | Anxiety (non-anxious/anxious) | 609/309 | 487/383 | 1096 (61%)/692 (39%) |
|  | Age (years) | 27.08 ± 8.12 | 26.70 ± 8.96 | 26.90 ± 8.54 |

Paired counts follow the category order shown in the row labels. Age is reported as mean ± SD.

### C. Speech Data Collection

Speech recordings were collected using the Sonde voice-recording application embedded within the web-based SurveyLex platform. Participants completed the recordings on their own devices, including smartphones, tablets, laptops, or desktop computers. Before recording, standardized on-screen instructions asked participants to use a quiet environment, minimize background sounds such as television or music, hold the device close to the mouth, and speak in a natural voice. Participants also completed a brief microphone check before proceeding. Audio files were stored in WAV format at a sampling rate of 48 kHz and converted to 16-bit PCM WAV format for analysis. Participants completed two neutral read-speech tasks: a short sentence (SS) and a long passage (LP). As summarized in Table II, the short sentence was designed to provide broad phonetic coverage [60], whereas the long passage provided an extended speech sample for acoustic feature extraction [61]. By design, these tasks were not intended to elicit anxiety or emotional arousal, but rather to support standardized speech collection across a large remote cohort without real-time clinical supervision. The data reflect practical conditions relevant to scalable voice-based screening because recordings were obtained using participants’ personal devices in everyday recording environments. Together, these tasks enabled evaluation of whether acoustic features from short-sentence and long-passage recordings could support classification of elevated generalized anxiety symptoms.

**TABLE 2.** Neutral read-speech tasks for acoustic feature extraction.

| **Task** | **Reading Text** | **Purpose** |
|---|---|---|
| Short sentence (SS) | The quick brown fox jumps over the lazy dog. | Comprehensive phonetic context |
| Long passage (LP) | John and I went to the farm in June. The sun shone all day, and the wind waved the grass in wide fields that ran by the road. Most birds had left on their track south, but old friends were there to greet us. Piles of wood had been stacked by the door, left there by the man who lives 12 miles down the road. | Acoustic feature extraction from extended speech |

### D. Speech Sample Preparation and Inclusion Criteria

Speech recordings were preprocessed and quality-screened before acoustic feature extraction. Voice activity detection (VAD) was first applied to each audio file using the WebRTC Python interface to remove prolonged non-speech intervals, particularly leading and trailing silences [62]. The VAD aggressiveness parameter was set to 1, a relatively conservative setting intended to trim silence while retaining the primary speech content. Each recording was then manually reviewed to confirm that it contained the intended speech task and a single analyzable repetition. When needed, recordings were trimmed to remove extra speech, repeated attempts, or other non-task material.

Recordings were excluded if they were incomplete, corresponded to the wrong task, contained multiple repetitions that could not be cleanly separated, had poor audio quality, or were affected by excessive background noise. Automated denoising was not applied because noise-reduction procedures can alter acoustic properties relevant to voice-based modeling, particularly frequency-, perturbation-, and spectral-domain features. Manual screening was therefore used to remove noisy data and preserve the integrity of the acoustic signal under remote recording conditions.

The number of eligible recordings differed between the SS and LP tasks because participants completed the speech tasks remotely and not all recordings passed quality screening. Some participants contributed valid recordings for both tasks, whereas others contributed only one eligible speech sample. Missingness in the GAD-7 questionnaire was handled consistently with the earlier analysis of this cohort: when one or two items were missing, the missing responses were replaced with the mode of the participant’s available responses; participants with more than two missing GAD-7 items or missing demographic information were excluded. The complete country-specific recruitment, speech-quality screening, and missingness-handling flow chart is summarized in Fig. 1.

**Fig. 1.**
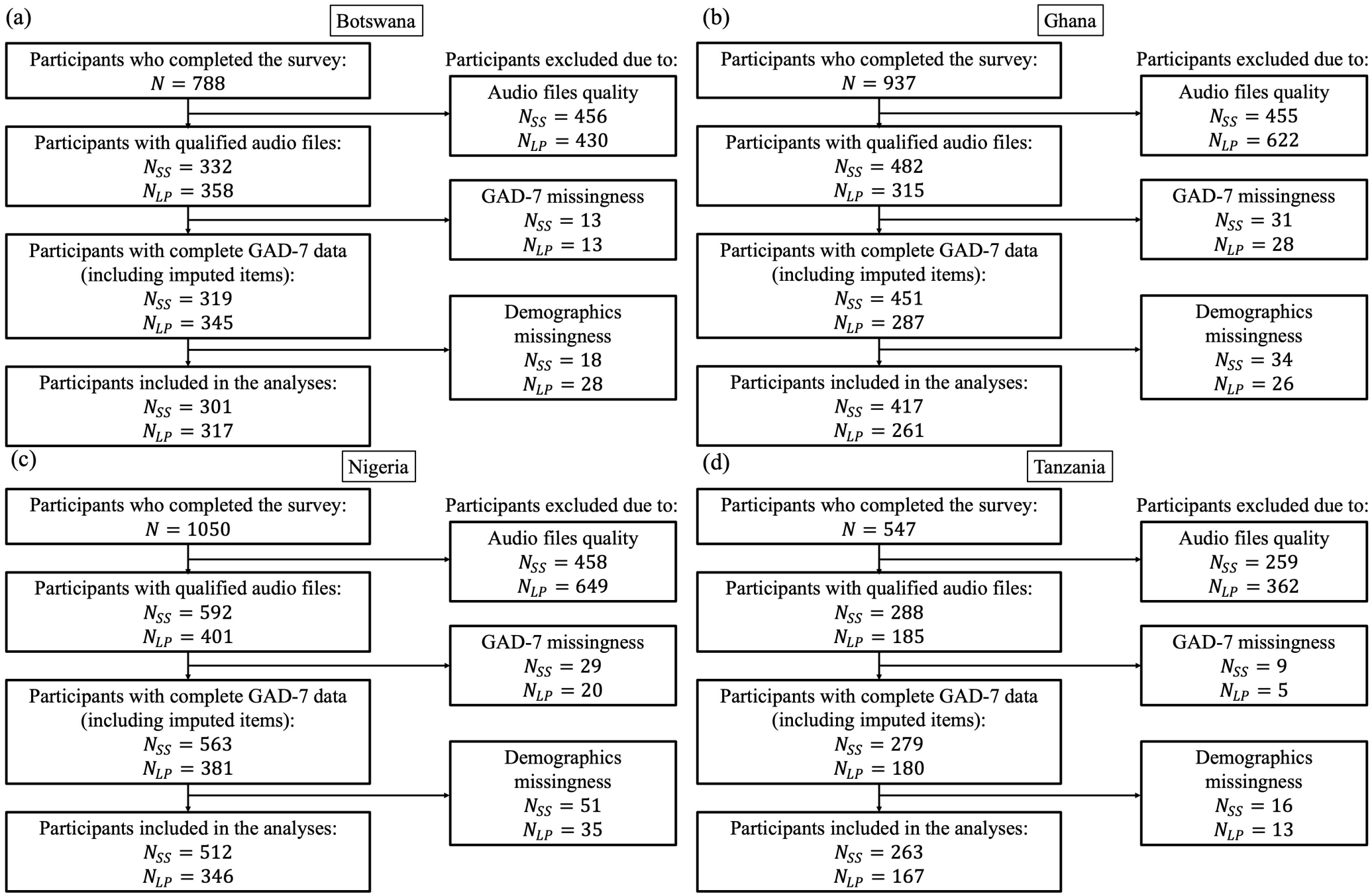
Participant screening and inclusion by country. *N*_*SS*_ and *N*_*LP*_ denote the numbers of SS and LP recordings, respectively, at each stage.

### E. Feature Extraction

The eGeMAPS acoustic feature set (version v02) [48] was extracted separately from each eligible SS and LP recording using the Python interface of openSMILE [63]. This standardized, interpretable representation comprises 88 recording-level features derived from 25 LLDs, capturing frequency, amplitude, spectral, cepstral, and temporal properties. It extends our earlier fundamental frequency (F_0_) focused analysis of the same cohort [20] by capturing a broader range of potentially anxiety-relevant acoustic properties. Two feature configurations were evaluated. The primary acoustic-only configuration included eGeMAPS features. The secondary context-augmented configuration additionally included age and binary self-reported health status in all modeling scenarios, and binary sex in combined-sex scenarios.

### F. Machine Learning Pipeline

A machine learning pipeline was developed primarily to evaluate screening-level classification of elevated generalized anxiety symptoms. All pipeline steps, hyperparameter tuning, validation, and model-interpretation analyses were implemented in Python. The main software packages and versions used in the analysis are reported in Supplementary Table S1. Nine modeling scenarios were constructed by crossing speechtask configuration (SS, LP, or combined-task) with sex stratification (male-only, female-only, or combined-sex), yielding scenarios denoted as D_1_–D_9_. Note that in combined-task scenarios, SS and LP recordings were pooled as separate observations. Then, for each scenario, classifiers were trained using both acoustic-only and context-augmented feature configurations, as summarized in Table III.

**TABLE 3.** Modeling scenarios by speech task and sex stratification.

| **Notation** | **Description** |
|---|---|
| D<sub>1</sub> | SS & male |
| D<sub>2</sub> | LP & male |
| D<sub>3</sub> | Both tasks & male |
| D<sub>4</sub> | SS & female |
| D<sub>5</sub> | LP & female |
| D<sub>6</sub> | Both tasks & female |
| D<sub>7</sub> | SS & both sexes |
| D<sub>8</sub> | LP & both sexes |
| D<sub>9</sub> | Both tasks & both sexes |

All data-dependent preprocessing and model fitting were applied to training partitions within each CV fold to minimize leakage. Continuous acoustic features and age underwent Yeo–Johnson transformation to reduce skewness and standardization to zero mean and unit variance. Local outlier factor (LOF) identified training outliers through local density comparisons, and the synthetic minority oversampling technique (SMOTE) addressed class imbalance by generating synthetic minority-class samples. Optional recursive feature elimination (RFE) reduced dimensionality by iteratively removing less informative features, and parallel pipelines without RFE assessed its effect. For context-augmented models, categorical predictors were neither power-transformed nor standardized, and SMOTE was replaced with its variant for nominal and continuous features, SMOTENC, to handle categorical variables during resampling.

Classification used repeated nested CV, with 10-fold stratified group CV repeated 10 times in the outer loop and 10-fold stratified group CV in the inner loop. Participant-level grouping prevented recordings from the same individual from appearing in both training and held-out partitions within either the inner or outer CV loop, while stratification approximately preserved class balance. Within each outer training fold, randomized search evaluated 300 hyperparameter configurations using AUC-ROC as the objective. Across the 100 outer folds, this yielded 30,000 hyperparameter trials, corresponding to 300,000 inner-fold pipeline fits per classifier, modeling scenario, feature configuration, and pipeline variant. The selected configuration was refitted on the full outer training fold and evaluated on the held-out fold. Search spaces and fixed hyperparameters are provided in Supplementary Tables S2–S3. Classifiers included support vector classifier (SVC), decision tree (DT), random forest (RF), gradient boosting (GB), extreme gradient boosting (XGB), light gradient boosting machine (LGBM), multilayer perceptron (MLP), Gaussian naive Bayes (NB), and k-nearest neighbors (KNN).

Model performance was assessed on the held-out outer folds using AUC-ROC, balanced accuracy, average precision, and F1 score. AUC-ROC was used as the primary metric because it evaluates discrimination across decision thresholds and is appropriate for imbalanced binary classification. Balanced accuracy, also referred to as unweighted average recall (UAR), was reported to account for unequal class sizes. Average precision was used to summarize precision– recall performance, which is particularly informative when the positive class is less prevalent. F1 score was also reported as a complementary threshold-dependent measure combining precision and recall. SHAP values were computed for for sampled held-out recordings and averaged per recording across repeated outer-fold evaluations. Then, features were ranked by the mean absolute value of these averaged contributions.

Secondary regression analyses predicted continuous GAD-7 scores using equivalent regressors, excluding NB because no directly corresponding regression estimator was available.

These analyses retained the same modeling scenarios, feature configurations, and fold-specific preprocessing, but used GroupKFold in both CV loops and omitted class-imbalance resampling. Hyperparameters were selected by minimizing root mean squared error (RMSE), using search spaces analogous to those in Supplementary Tables S2–S3. RMSE was the primary evaluation metric, complemented by mean absolute error (MAE) and the coefficient of determination (*R*^2^).

## III. RESULTS

This section reports model performance across the nine modeling scenarios (see Table III), two feature configurations (acoustic-only and context-augmented), and two pipeline variants (with and without feature selection). Figure 2 first summarizes the mean AUC-ROC values for all machine learning models when feature selection was included in the pipeline across repeated outer folds. In the acoustic-only setting, models trained on eGeMAPS features showed limited discrimination: male-only scenarios remained close to chance level, whereas female-only and combined-sex scenarios showed modestly higher performance. No acoustic-only result exceeded an AUC-ROC of 0.60; the highest value was obtained by the RF model in the combined-sex long-passage scenario, D_8_, with an AUC-ROC of 0.594. Adding participant-level context improved performance relative to the corresponding acoustic-only models. The strongest result was obtained by the XGB model in the female-only combined-task scenario, D_6_, with an AUC-ROC of 0.699, indicating moderate discrimination.

**Fig. 2.**
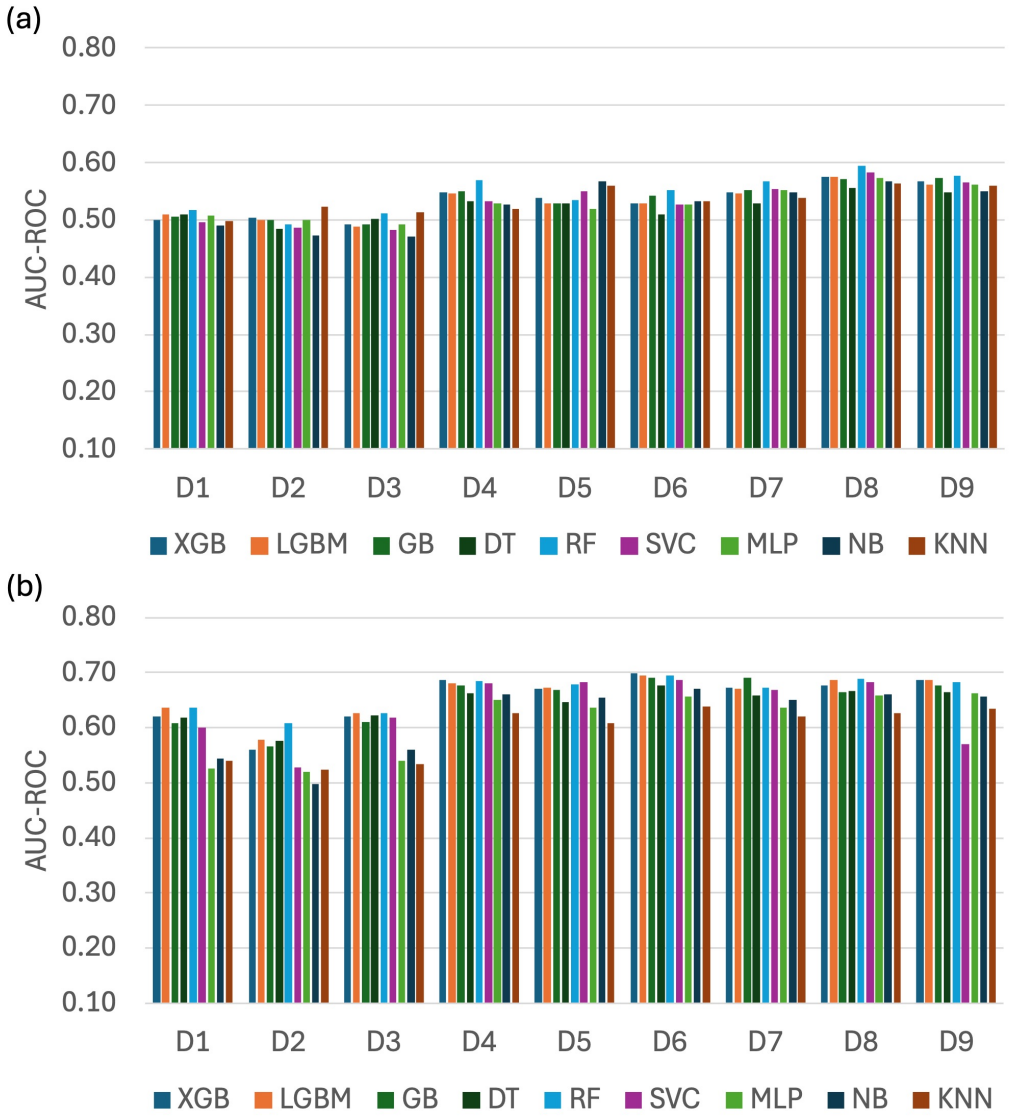
Mean AUC-ROC across repeated outer folds by modeling scenario for each ML model, using (a) acoustic-only and (b) context-augmented features with RFE.

Additional performance metrics, including balanced accuracy, average precision, and F1 score, are summarized in Figs. S1–S3. Overall, balanced accuracy followed a pattern similar to that of AUC-ROC but with lower absolute values, supporting the same relative trends in modeling scenarios while accounting for class imbalance. Average precision was generally higher in the context-augmented setting than in the acoustic-only setting, indicating improved precision–recall performance for the elevated generalized anxiety symptom class. The F1 score showed a similar overall pattern, with higher values in the context-augmented setting than in the acoustic-only setting.

Feature selection had a model-dependent effect, with no consistent improvement across modeling scenarios and feature configurations (see Fig. S4). In the acoustic-only setting, RF achieved nearly identical AUC-ROC values in scenario D_8_ with and without RFE (0.594 ± 0.049 and 0.596 ± 0.049, respectively). In the context-augmented setting, on the other hand, the highest AUC-ROC values in scenario D_6_ were obtained by XGB with RFE (0.699 ± 0.056) and LGBM without RFE (0.692 ± 0.052). Notably, error bars represent standard deviations across repeated outer folds, illustrating variability across training and validation partitions and underscoring the importance of evaluating performance beyond a single split or CV run.

Scenario-level performance, summarized by averaging AUC-ROC across classifiers (see Table IV), varied with sex stratification and speech-task configuration. Context-augmented models consistently outperformed their acoustic-only counterparts across all nine scenarios. Acoustic-only performance was highest in combined-sex scenarios (D_7_–D_9_), whereas female-only and combined-sex scenarios (D_4_–D_9_) showed the strongest context-augmented performance. Male-only scenarios (D_1_–D_3_) yielded the lowest averages in both settings, representing the most consistent difference across scenario groups. Within combined-sex scenarios, LP recordings outperformed SS and combined-task recordings under both feature configurations. However, no consistent speech-task advantage emerged within male-only or female-only scenarios, indicating that the LP advantage was specific to the combined-sex analyses.

**TABLE 4.** AUC-ROC by modeling scenario for considered feature sets with RFE (mean *±* SD across classifiers).

| **Scenario** | **Acoustic-only** | **Context-augmented** |
|---|---|---|
| D<sub>1</sub> | 0.504 ± 0.008 | 0.591 ± 0.044 |
| D<sub>2</sub> | 0.496 ± 0.014 | 0.551 ± 0.035 |
| D<sub>3</sub> | 0.494 ± 0.014 | 0.595 ± 0.039 |
| D<sub>4</sub> | 0.539 ± 0.016 | 0.668 ± 0.020 |
| D<sub>5</sub> | 0.540 ± 0.016 | 0.657 ± 0.024 |
| D<sub>6</sub> | 0.531 ± 0.011 | **0.678 ± 0.021** |
| D<sub>7</sub> | 0.548 ± 0.011 | 0.660 ± 0.021 |
| D<sub>8</sub> | 0.573 ± 0.011 | 0.668 ± 0.019 |
| D<sub>9</sub> | 0.563 ± 0.010 | 0.658 ± 0.037 |

Model-level performance, summarized by averaging AUC-ROC across modeling scenarios (Table V), showed limited variation among acoustic-only classifiers, with means ranging from 0.523 to 0.546. Adding participant-level context improved average performance across all classifiers and produced greater differences between models. RF achieved the highest context-augmented mean AUC-ROC (0.663 ± 0.032), followed closely by LGBM, XGB, and GB. These averages highlight the stronger performance of tree-based ensembles in the context-augmented setting, whereas classifier choice had comparatively little influence when predictions relied on acoustic features alone. Furthermore, the corresponding scenario-and model-level summaries for the considered feature sets without RFE are provided in the Supplementary Tables S4 and S5, respectively.

**TABLE 5.** AUC-ROC by classifier for considered feature sets with RFE (mean *±* SD across scenarios).

| **Model** | **Acoustic-only** | **Context-augmented** |
|---|---|---|
| XGB | 0.534 ± 0.030 | **0.654 ± 0.045** |
| LGBM | 0.532 ± 0.029 | **0.659 ± 0.039** |
| GB | 0.535 ± 0.030 | 0.650 ± 0.044 |
| DT | 0.523 ± 0.023 | 0.643 ± 0.032 |
| RF | 0.546 ± 0.034 | **0.663 ± 0.032** |
| SVC | 0.531 ± 0.036 | 0.635 ± 0.059 |
| MLP | 0.529 ± 0.028 | 0.609 ± 0.062 |
| NB | 0.525 ± 0.038 | 0.617 ± 0.065 |
| KNN | 0.534 ± 0.023 | 0.594 ± 0.048 |

As a secondary analysis, the corresponding regression models were evaluated for continuous prediction of GAD-7 scores. Across modeling scenarios and feature configurations with feature selection, RMSE values were approximately 4–5.5 and MAE values approximately 3–4.5 (see Supplementary Fig. S5). In the illustrative example of RF regression performance for scenario D_9_, *R*^2^ values were 0.02 and 0.13 for the acoustic-only and context-augmented models, respectively. Predictions were concentrated largely below 10, overestimating lower observed scores and underestimating higher scores (see Supplementary Fig. S6). These findings indicate limited ability to capture variation in symptom severity.

Figures 3 and 4 present the top 10 features ranked by mean absolute SHAP value after averaging signed SHAP values for each recording across its sampled outer-fold evaluations, for RF, LGBM, and XGB in scenario D_9_ with feature selection. These classifiers were selected based on their highest mean context-augmented AUC-ROC across modeling scenarios (see Table V). In the acoustic-only setting, feature contributions were distributed across multiple acoustic descriptors, with no single feature dominating the rankings. Across RF, LGBM, and XGB, F_0_-related features appeared most consistently among the top-ranked predictors, particularly the mean and percentile values of semitone-scaled fundamental frequency. Other features, including equivalent sound level, formantrelated descriptors, voiced-segment measures, spectral flux, and slope-related features, also appeared among the top 10, although their rankings varied across models.

**Fig. 3.**
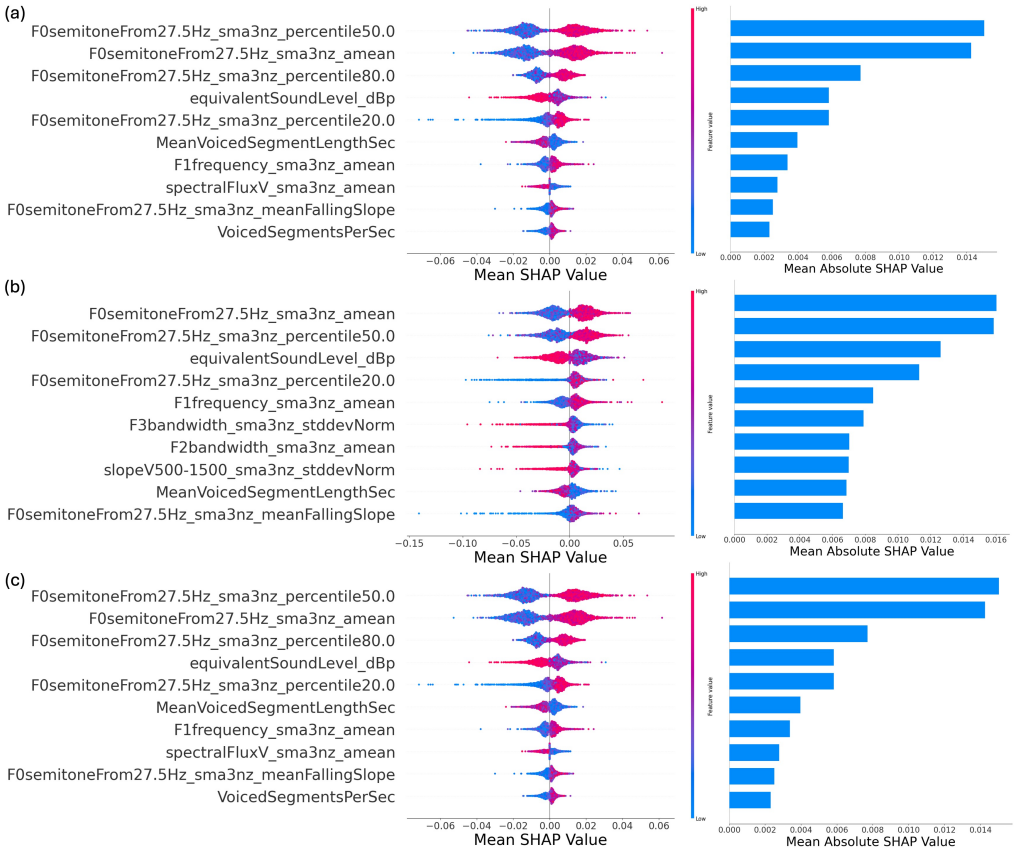
Top 10 acoustic features from models trained with RFE for modeling scenario D_9_: (a) RF, (b) LGBM, and (c) XGB. Features are ranked by mean absolute SHAP values after averaging per recording. Left: signed SHAP distributions; right: mean absolute SHAP values.

**Fig. 4.**
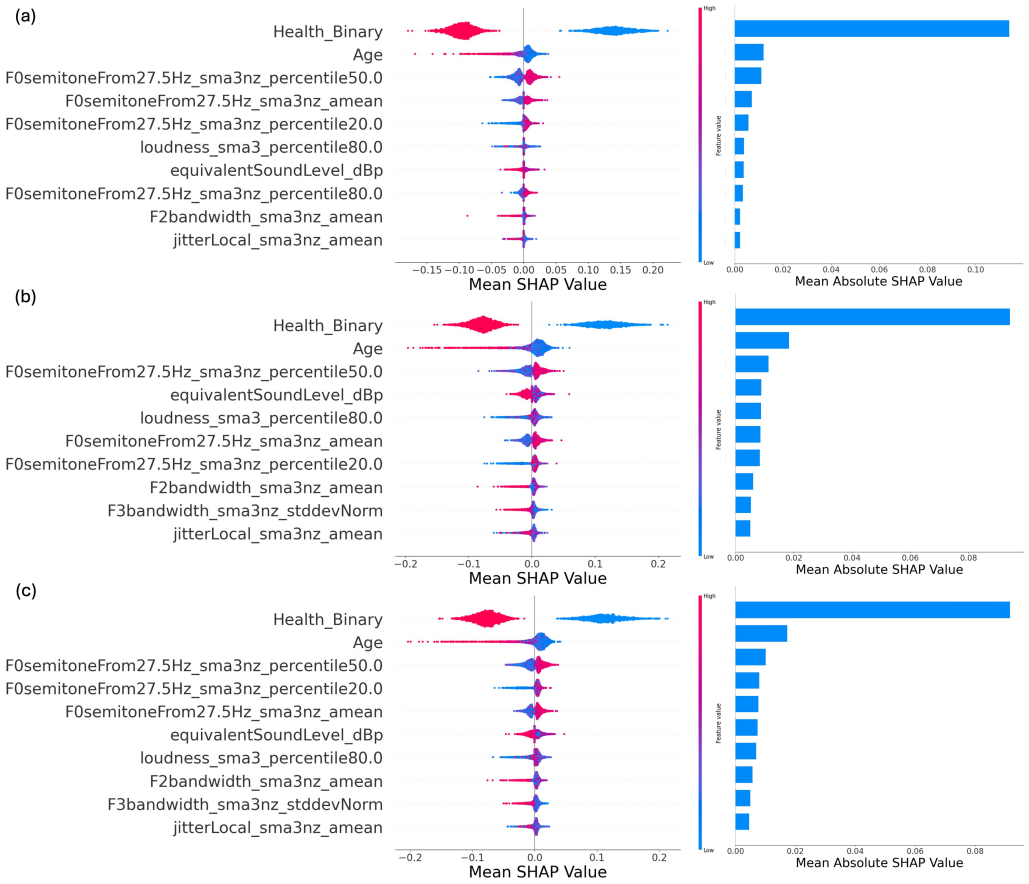
Top 10 predictors from context-augmented models trained with RFE for modeling scenario D9: (a) RF, (b) LGBM, and (c) XGB. Features are ranked by mean absolute SHAP values after averaging per recording. Left: signed SHAP distributions; right: mean absolute SHAP values.

In the context-augmented setting, the SHAP rankings changed markedly, with participant-level context variables showing the largest mean absolute SHAP values. Self-reported health status consistently had the largest contribution across the three models, followed by age, whereas binary sex did not appear among the top-ranked features. Acoustic descriptors remained represented among the top 10 features, including F_0_-related, loudness-related, formant-bandwidth, and jitter-related measures. However, their mean absolute SHAP values were smaller than those of health status and age, indicating smaller, model-dependent acoustic contributions to predictions within the context-augmented models. It should be noted that since the SPL of the recordings were not calibrated, the SHAP contributions of loudness-related and sound-level features should be interpreted cautiously, as these measures may reflect differences in microphone sensitivity, recording gain, and mouth-to-microphone distance in addition to vocal loudness.

## IV. DISCUSSION

This study evaluated whether remotely collected neutral read speech could support screening for elevated generalized anxiety symptoms. Acoustic-only models showed weak-to-modest discrimination, while adding demographic and health information improved performance, highlighting the importance of participant context in interpreting voice-based screening results.

In the acoustic-only analysis, the strongest performance was observed in the combined-sex long passage scenario, D_8_, where the RF model achieved similar AUC-ROC values with and without feature selection (0.594 ± 0.049 and 0.596 ± 0.049, respectively). These results indicate weak-to-modest discrimination and should be interpreted in light of the study design as recordings were remotely collected from neutral read-speech tasks and were not intended to elicit anxiety or emotional arousal. In contrast, studies reporting higher discrimination have often used anxiety-eliciting tasks or focused on specific anxiety subtypes, particularly social anxiety disorder (SAD). For example, Ciampelli et al. [45] reported AUC-ROC values of 0.854 and 0.739 for longitudinal SAD classification in males and females, respectively, using RF models trained on eGeMAPS features extracted from speech collected during the Trier Social Stress Test (TSST). In a more comparable audio-only setting, Brueckner et al. [56] used spontaneous speech to classify GAD-7-defined anxiety status and reported a UAR of 0.56 with an SVM model using eGeMAPS features, while their best deep learning model using Wav2Vec2 representations reached a UAR of 0.60. Sousa et al. [46] similarly analyzed smartphone-based free conversational speech for depression and anxiety screening in pregnant women and reported a best AUC-ROC of 0.71 using a combination of 64 MFCCs and eGeMAPS features (extracted by openSMILE) with a Light-GBM classifier. Although these studies differ in speech task, label definition, feature representation, and evaluation metric, they collectively suggest that identifying generalized anxiety symptoms from non-eliciting, remotely collected speech remains challenging, particularly when relying on compact handcrafted acoustic descriptors alone. Although methodological differences limit direct comparison, the reported modest performance, especially by Brueckner et al. [56] in a study setting comparable to ours, broadly aligns with our acoustic-only findings. Our study extends this evidence to remotely collected neutral read speech in a large, multi-country African cohort, further highlighting the limited discrimination achieved with eGeMAPS features alone.

Speech tasks also influenced performance, although the effect was scenario-dependent. In the combined-sex scenarios, the LP task consistently outperformed both the SS and combined tasks in the acoustic-only and context-augmented settings with RFE (see Table IV). This pattern suggests that longer read-speech recordings may provide more stable estimates of eGeMAPS statistical functionals, particularly in larger and more heterogeneous samples. However, this advantage was not consistent in the single-sex scenarios, indicating that the benefit of longer recordings depended on the modeling population. This interpretation is consistent with Tasnim et al. [43], who reported lower prediction errors for longer and more linguistically rich speech tasks, such as picture description and prompted narrative, than for shorter phoneme-based tasks. Although standardized English texts reduced variation in linguistic content, differences in participants’ language backgrounds and English proficiency in the present study, may have influenced prosodic and temporal characteristics, complicating the interpretation of task-related performance differences.

Sex-related differences also shaped classification performance. Female-only scenarios generally outperformed male-only scenarios across both feature settings, although this does not establish that anxiety-related acoustic patterns were intrinsically more detectable in females. Possible sex-related differences in symptom reporting could also have influenced these comparisons, as classification relied on self-reported GAD-7 labels. In the acoustic-only setting, combined-sex scenarios showed a slight advantage over female-only scenarios, possibly reflecting the benefit of larger sample size and broader acoustic variability. In the context-augmented setting, combined-sex models remained competitive with female-only models, although this comparison should be interpreted cautiously because sex was included as a predictor in the combined-sex scenarios. These findings indicate that sex may influence both anxiety-related acoustic markers and model behavior. Prior studies provide related but not identical evidence. Ciampelli et al. [45] reported sex-dependent acoustic predictors and stronger SAD classification in males than females using speech elicited during the TSST, whereas Alon-Ronen et al. [28] found that sex-specific classifiers outperformed a sex-unified configuration for social anxiety classification. These findings suggest that the value of sex stratification depends on the speech task, anxiety construct, sample characteristics, and feature configuration.

The context-augmented analysis evaluated whether adding participant-level demographic and health-related variables to eGeMAPS features improved speech-based classification of elevated generalized anxiety symptoms. These models consistently outperformed acoustic-only models in scenario-level and model-level summaries (see Tables IV and V). Performance was strongest in the female-only combined-task scenario, D_6_, with AUC-ROC values of 0.699 ± 0.056 for XGB with feature selection and 0.692 ± 0.052 for LGBM without feature selection, indicating moderate discrimination. SHAP analysis identified self-reported health status and age as the largest contributors to model predictions in the combined-sex, combinedtask scenario, D_9_ (see Fig. 4), whereas acoustic descriptors contributed more modestly and varied across models. The present analysis extends our earlier F_0_-focused study [20] by highlighting several F_0_-related descriptors among the leading acoustic predictors across models in both feature settings, whereas F_0_-min was the most prominent acoustic contributor previously. The current semitone-scaled eGeMAPS descriptors summarize a broader range of F_0_ characteristics than the earlier Hz-based measures, although their contributions remained smaller than those of health status and age in context-augmented models. Additionally, the importance of participant-level context is also consistent with Teferra et al. [27], who reported that GAD screening performance increased from an AUC-ROC of 0.57 using acoustic and linguistic features alone to 0.62 after adding demographic variables. Similarly, Park et al. [29] found that combining acoustic eGeMAPS features with physiological measures outperformed single-source models for predicting severe anxiety symptoms in SAD.

The secondary regression analysis provided a complementary perspective on the distinction between screening and symptom-severity estimation. The regressors did not adequately capture continuous variation across the full GAD-7 range, with predictions concentrated largely between 0 and 10. This restricted prediction range and the systematic underestimation of higher observed scores indicate limited ability to estimate symptom severity, despite the modest discrimination observed in the separate classification analysis.

Model choice also contributed to performance differences, although its effect was smaller than that of feature configuration. As summarized in Table V, tree-based models, including RF and boosting-based classifiers such as XGB, LGBM, and GB, showed the most consistent performance in the context-augmented setting. This pattern is consistent with prior speech-based mental health studies in which ensemble and boosted models have been effective for capturing nonlinear relationships among acoustic and participant-level predictors. In contrast, classifier differences were relatively small in the acoustic-only setting, and all models remained within a weak discrimination range. Thus, although model selection contributed to performance variability, the available predictor information, particularly the contrast between acoustic-only and context-augmented feature sets, had a larger influence on classification performance than the choice of classifier alone.

Lastly, it is worth emphasizing that the reported performance metrics must be interpreted in light of the rigorous ML pipeline developed to reduce data leakage and overly optimistic estimation. Data-dependent preprocessing, outlier detection, resampling, and feature selection were confined to training folds, while participant-level grouping prevented recordings from the same individual from appearing in both training and held-out CV partitions. Hyperparameter tuning was restricted to the inner CV loop, separating model selection from outer-fold performance assessment. Repeated outer evaluations characterized variability across data partitions, and complementary metrics supported interpretation under class imbalance. These choices are central to assessing speech-based screening models because violations can inflate apparent discrimination without improving generalization.

## V. CONCLUSIONS

This study evaluated ML models for screening elevated generalized anxiety symptoms from remotely collected neutral read speech in a large, multi-country African cohort. Acoustic-only eGeMAPS features showed weak-to-modest discrimination, while adding demographic and health information consistently improved performance. SHAP analyses identified health status and age as the leading contributors in context-augmented models, with more modest, model-dependent acoustic contributions. Secondary regression was not able to capture continuous GAD-7 scores, with predictions concentrated in the low-to-mild range, indicating limited suitability for precise symptom-severity estimation. Performance varied with speechtask configuration, sex stratification, classifier, and feature processing, underscoring the importance of task design and rigorous evaluation. These findings support further investigation of voice-derived features alongside participant-level information for scalable remote anxiety screening. External validation, richer speech tasks, longitudinal data, and multimodal integration are needed to clarify their utility.

## Data Availability

The voice recordings are considered identifiable biometric data and cannot be shared. The additional data supporting the findings of this study are available from the corresponding author upon reasonable request.

## ACKNOWLEDGMENTS

This work was supported by the Tetrad Initiative for Interdisciplinary Research Funding through the Spartan Tetrad Program at Michigan State University and the National Research Foundation of South Africa, with additional support through computational resources and services provided by the Institute for Cyber-Enabled Research at Michigan State University. The authors acknowledge the use of generative artificial intelligence (ChatGPT 5.2 and 6.1 Sol) for code review and development and language editing and polishing. All content was directly reviewed, edited, and verified by the authors, who take full responsibility for the final published work.

