## Supplementary Materials for "Automated Screening for Generalized Anxiety Disorder Using Voice Analysis and Machine Learning"

Table S1. Python packages and their versions used for model development.

| **Python Package** | **Version** |
| --- | --- |
| numpy | 2.5.0 |
| pandas | 3.0.3 |
| scipy | 1.18.0 |
| matplotlib | 3.11.0 |
| scikit_learn | 1.9.0 |
| imbalanced_learn | 0.13.0 |
| lightgbm | 4.6.0 |
| xgboost | 3.3.0 |
| shap | 0.50.0 |

Table S2. Hyperparameters of pipeline steps.

| **Step** | **Hyperparameters** | **Value/Range** |
| --- | --- | --- |
| Power Transformation | method  standardize | Yeo-Johnson  True |
| Oversampling | SMOTE - k_neighbors  SMOTENC - k_neighbors | randint(3, 8)^*^  randint(3, 8) |
| Outlier Detection (LOF) | contamination  n_neighbors  algorithm  metric | 0.05  20  auto  manhattan |
| Feature Selection (RFE) | n_features_to_select  estimator  step | uniform(0.1, 0.9)  RandomForestClassifier(n_estimators=25)^**^  0.1 |

^**^ SciPy notation is followed: uniform(a, b) spans [a, a + b); randint(a, b) spans integers a to b − 1; loguniform(a, b) is uniform in log space between abounds a and b.

^*^ The regression counterpart was used for regression analyses

Table S3. Fixed hyperparameter values and search ranges for each model.

| **Model** | **Hyperparameters** | **Range** |
| --- | --- | --- |
| XGB | booster  tree_method  n_estimators  learning_rate  min_child_weight  max_depth  gamma  subsample  colsample_bytree  reg_lambda | gbtree  hist  randint(200, 1001)  loguniform(1e-3, 3e-1)  uniform(1, 7)  randint(3, 11)  uniform(0, 1)  uniform(0.6, 0.4)  uniform(0.6, 0.4)  loguniform(1e-3, 1e1) |
| LGBM | boosting_type  num_leaves  min_child_samples  max_depth  min_split_gain  n_estimators  learning_rate  colsample_bytree  reg_lambda | gbdt  randint(24, 91)  randint(15, 101)  randint(3, 11)  loguniform(1e-3, 1e1)  randint(200, 1001)  loguniform(1e-3, 3e-1)  uniform(0.6, 0.4)  loguniform(1e-3, 1e1) |
| GB | n_estimators  n_iter_no_change  tol  validation_fraction  learning_rate  max_depth  max_features  min_samples_split  min_samples_leaf  subsample | 1500  10  1e-4  0.10  loguniform(5e-3, 5e-1)  randint(3, 8)  ["sqrt", "log2", 0.3, 0.4, 0.5, 0.6, 0.7, 0.8, 0.9, 1]  uniform(0.05, 0.30)  uniform(0.01, 0.09)  uniform(0.75, 0.25) |
| DT | max_depth  max_features  min_samples_split  min_samples_leaf  ccp_alpha | randint(3, 21)  ["sqrt", "log2", 0.3, 0.4, 0.5, 0.6, 0.7, 0.8, 0.9, 1]  uniform(0.05, 0.35)  uniform(0.01, 0.09)  loguniform(1e-6, 1e-1) |
| RF | n_estimators  max_depth  max_features  min_samples_split  min_samples_leaf  ccp_alpha | randint(200, 1001)  randint(3, 21)  ["sqrt", "log2", 0.3, 0.4, 0.5, 0.6, 0.7, 0.8, 0.9, 1]  uniform(0.05, 0.45)  uniform(0.01, 0.19)  loguniform(1e-6, 1e-1) |
| SVC | C  gamma  kernel | loguniform(1e-3, 1e6)  loguniform(1e-6, 1e2)  ["rbf"] |
| MLP^*^ | max_iter  early_stopping  n_iter_no_change  validation_fraction  hidden_layer_counts  hidden_layer_widths  learning_rate_init  batch_size  activation  solver  alpha | 2000  True  15  0.15  [3, 4, 5, 6]  [16, 32, 64, 128]  loguniform(1e-5, 1e-2)  [16, 32, 64, 128, "auto"]  ["relu", "tanh", "logistic"]  ["adam", "sgd", "lbfgs"]  loguniform(1e-5, 1e-2) |
| NB | var_smoothing | loguniform(1e-11, 1e-7) |
| KNN | n_neighbors  weights  algorithm  p | list(range(3, 22, 2))  ["uniform", "distance"]  ["ball_tree", "kd_tree", "brute"]  [1, 2] |

^*^ hidden_layer_sizes was constructed from the selected layer count and width, with the same width applied to all hidden layers.

| 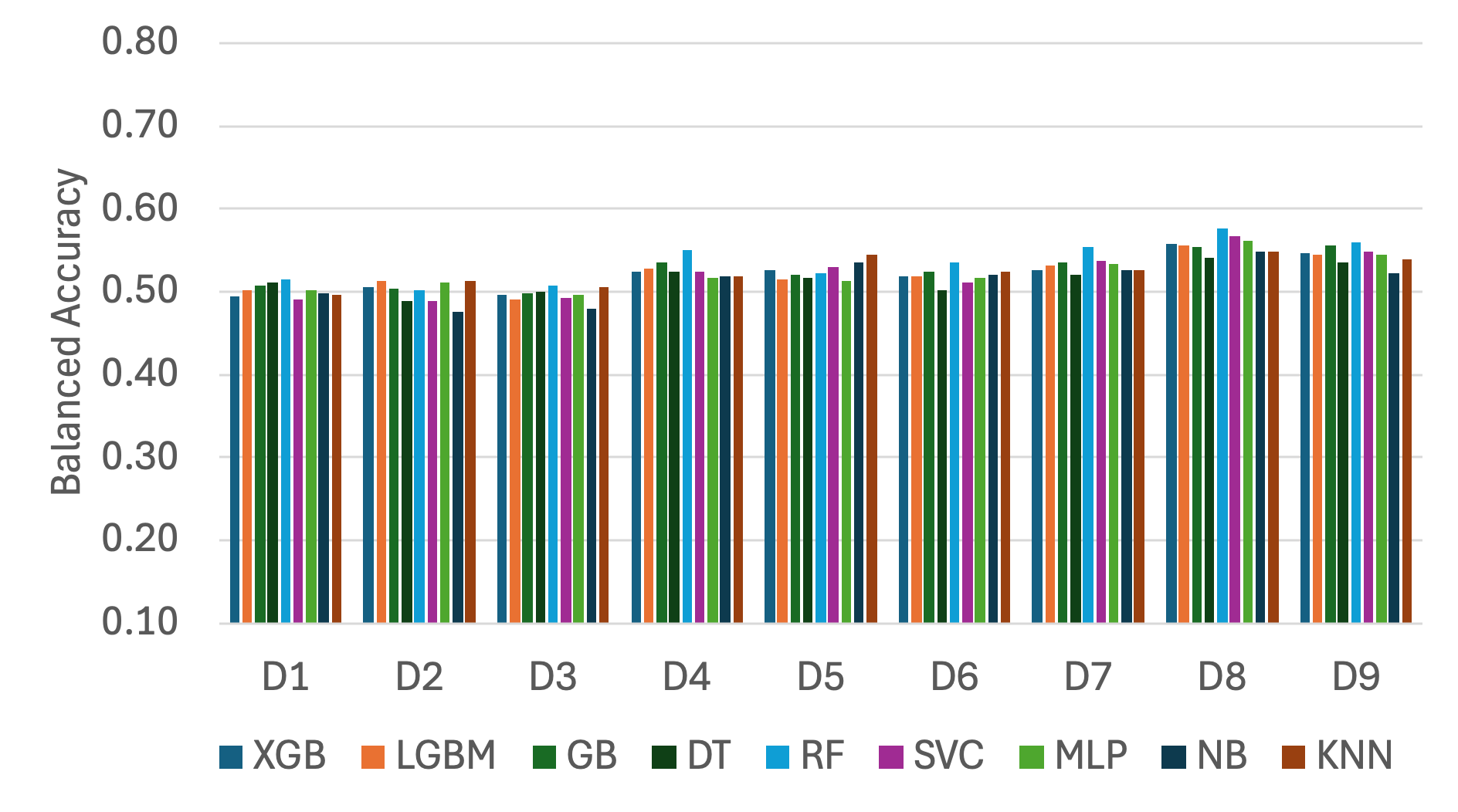 | 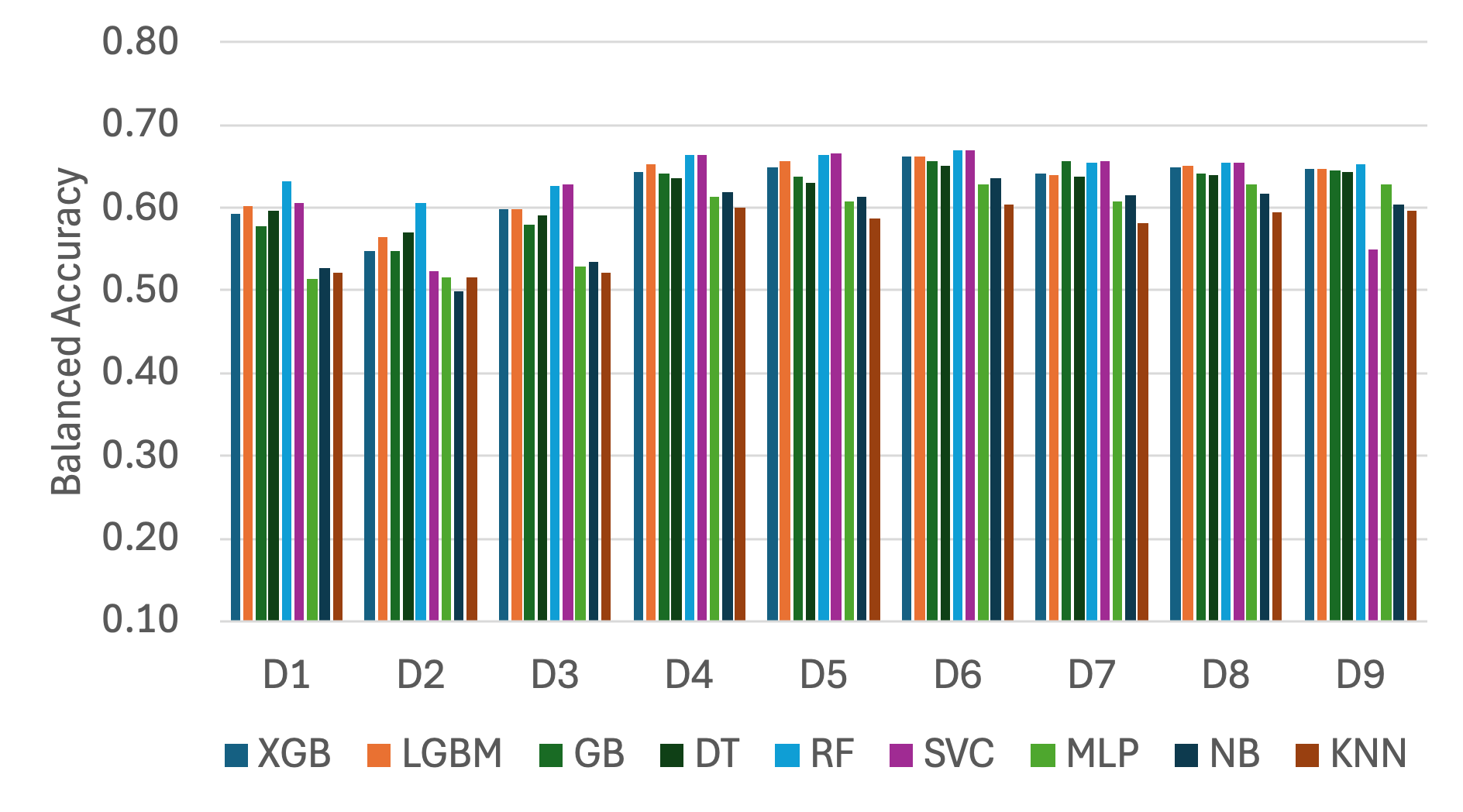 |
| --- | --- |

Fig. S1. Mean balanced accuracy across repeated outer folds by modeling scenario for each ML model, using (a) acoustic-only and (b) context-augmented features with RFE.

(a)

(b)

(b)

(a)

| 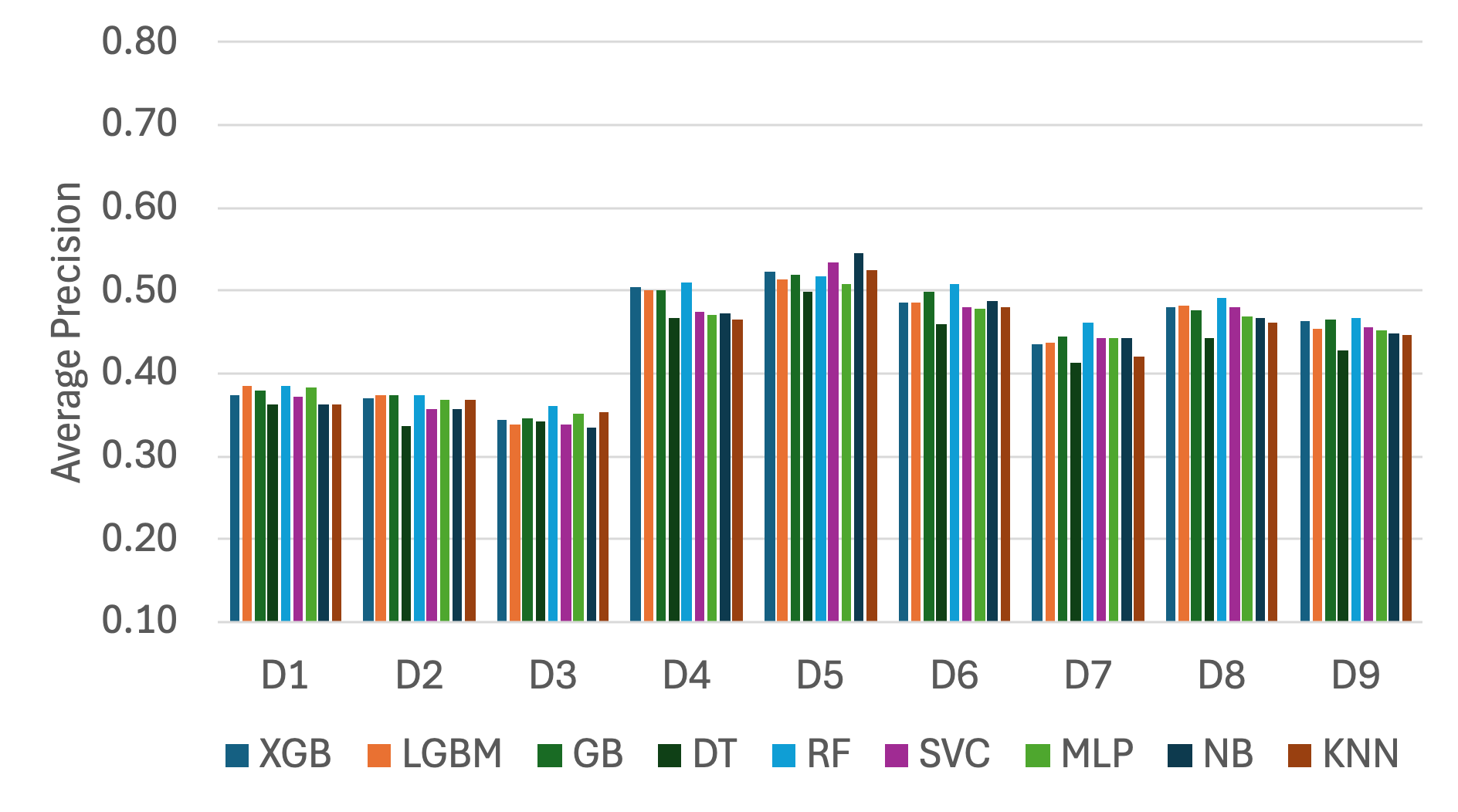 | 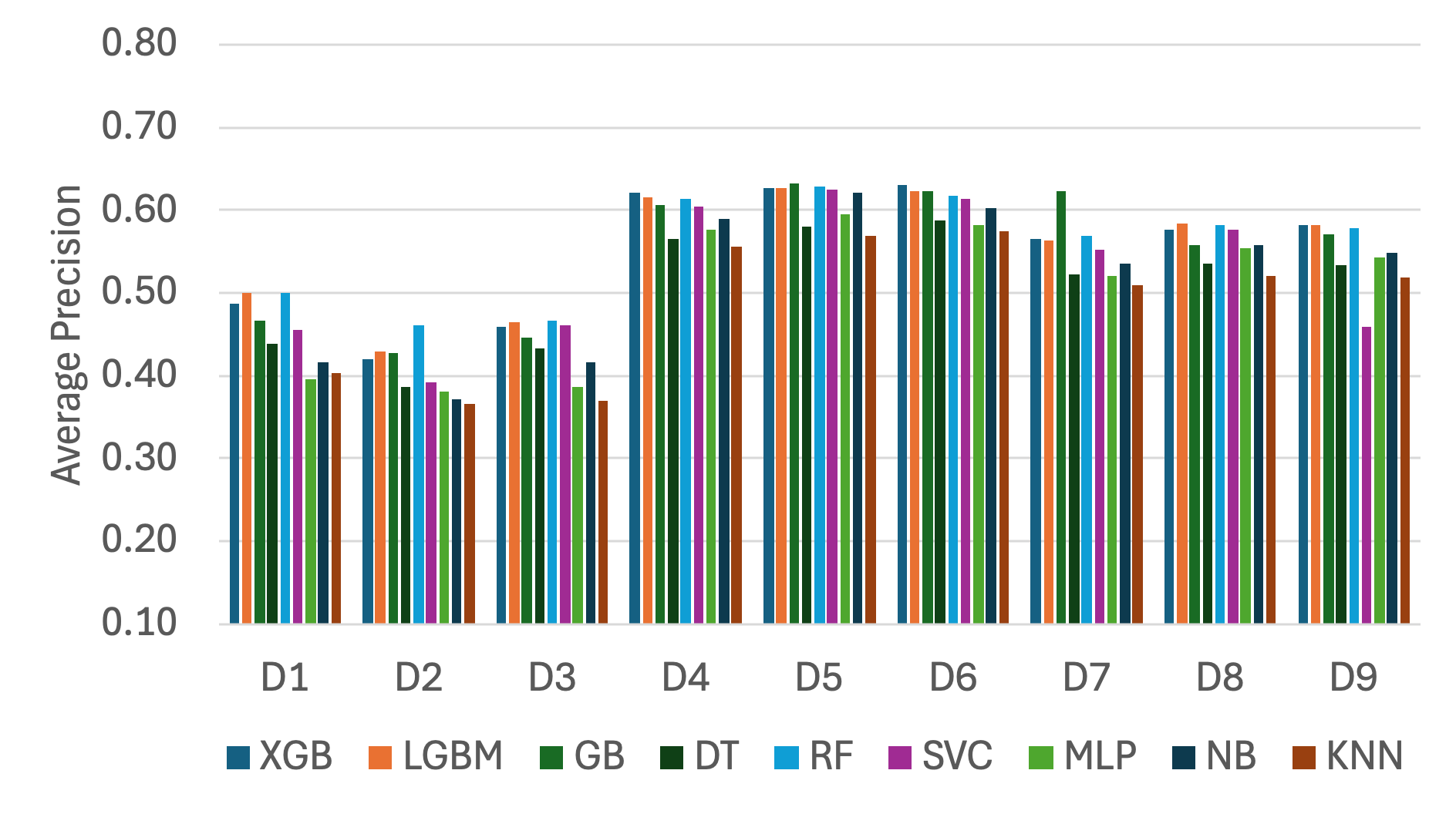 |
| --- | --- |

Fig. S2. Mean average precision across repeated outer folds by modeling scenario for each ML model, using (a) acoustic-only and (b) context-augmented features with RFE.

(a)

(b)

| 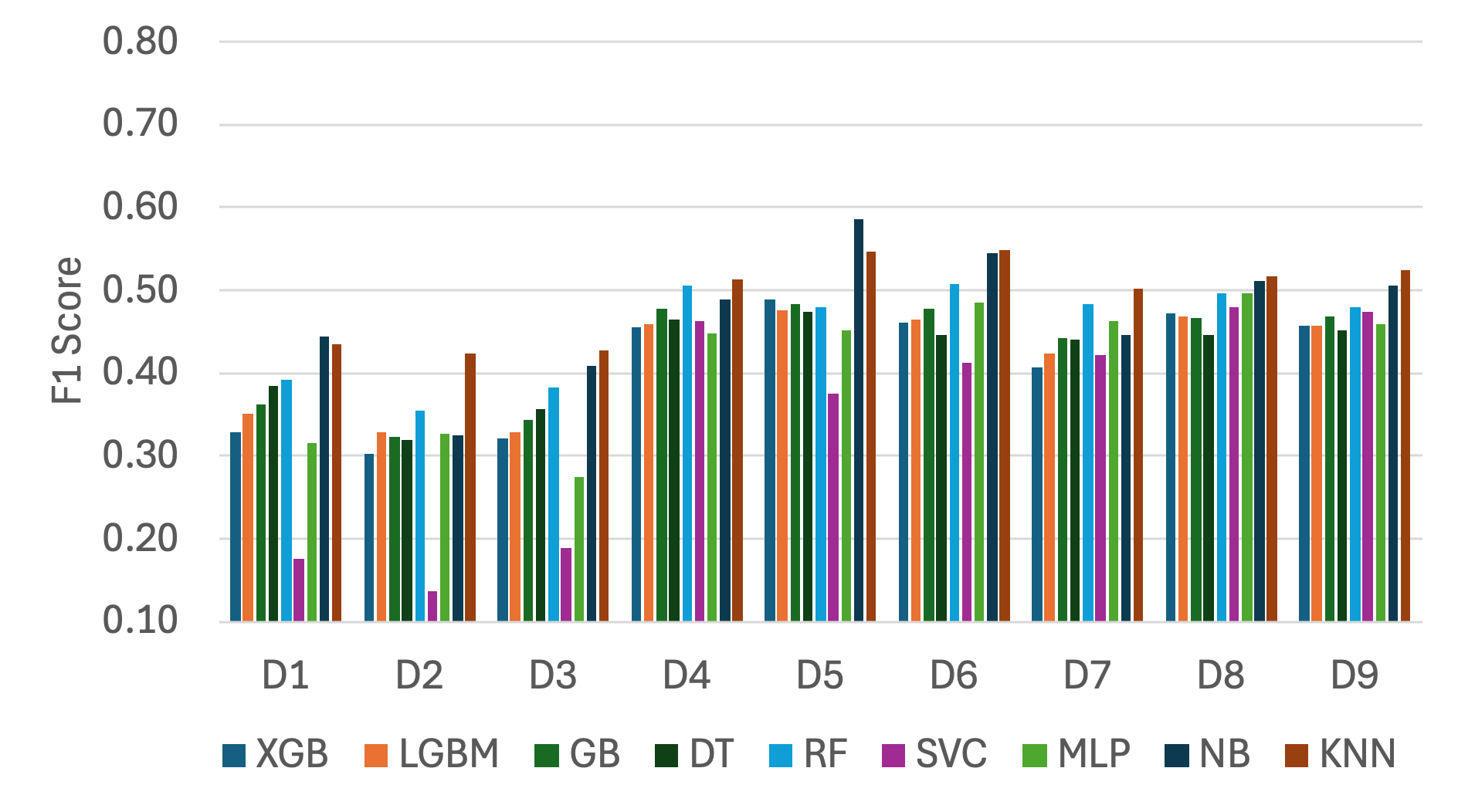 | 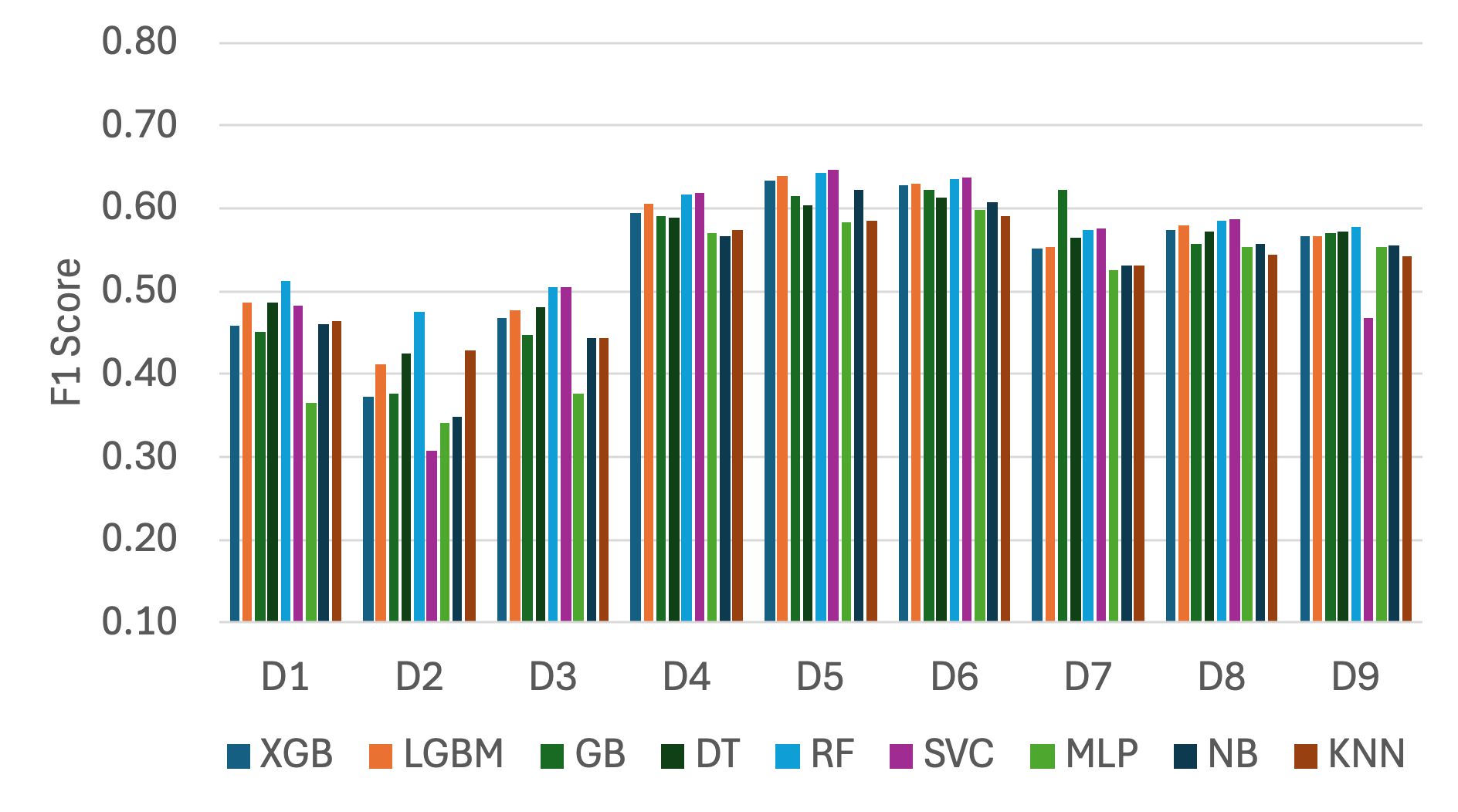 |
| --- | --- |

Fig. S3. Mean F1 score across repeated outer folds by modeling scenario for each ML model, using (a) acoustic-only and (b) context-augmented features with RFE.

| Acoustic-only | Context-augmented | Acoustic-only | Context-augmented |
| --- | --- | --- | --- |
| 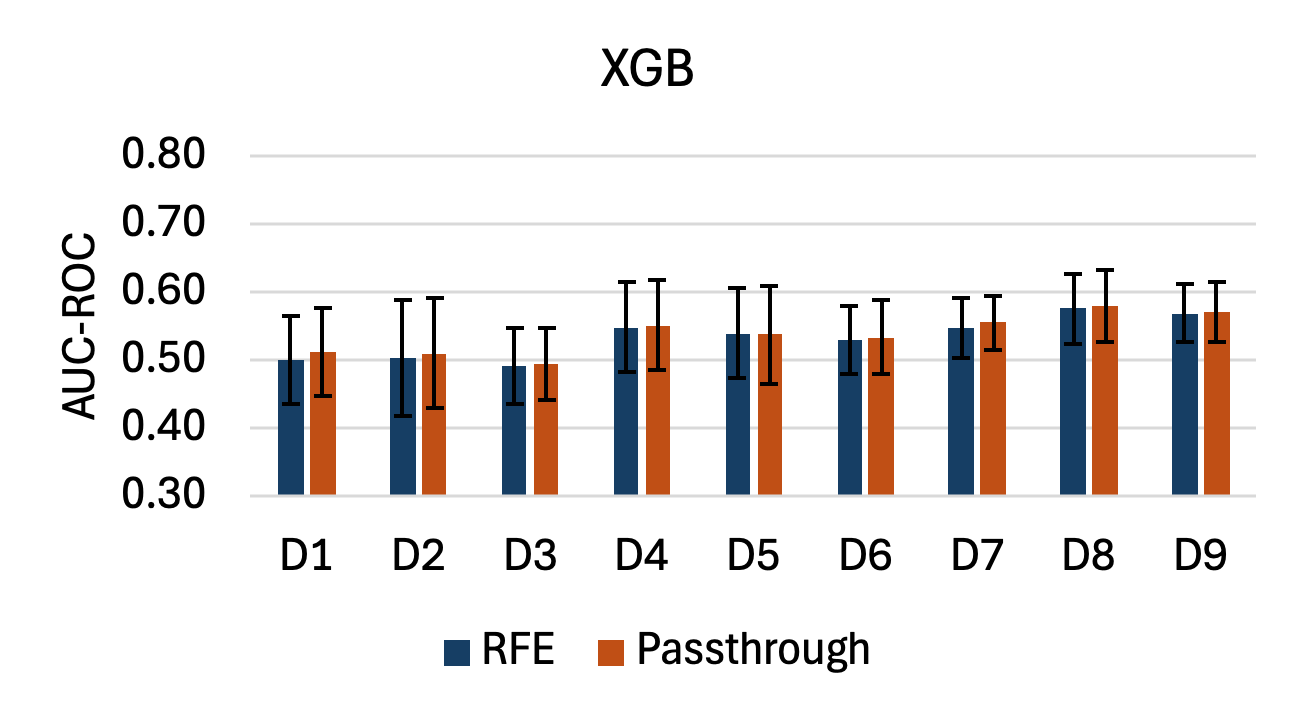 | 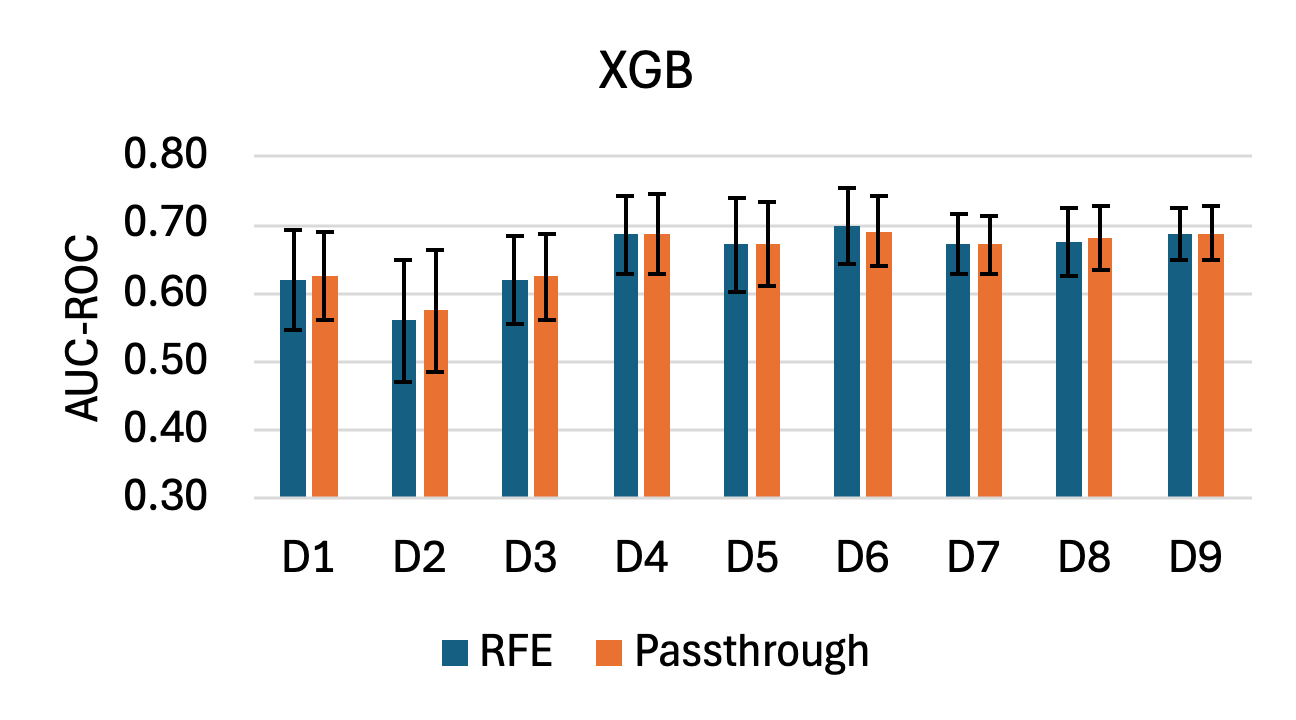 | 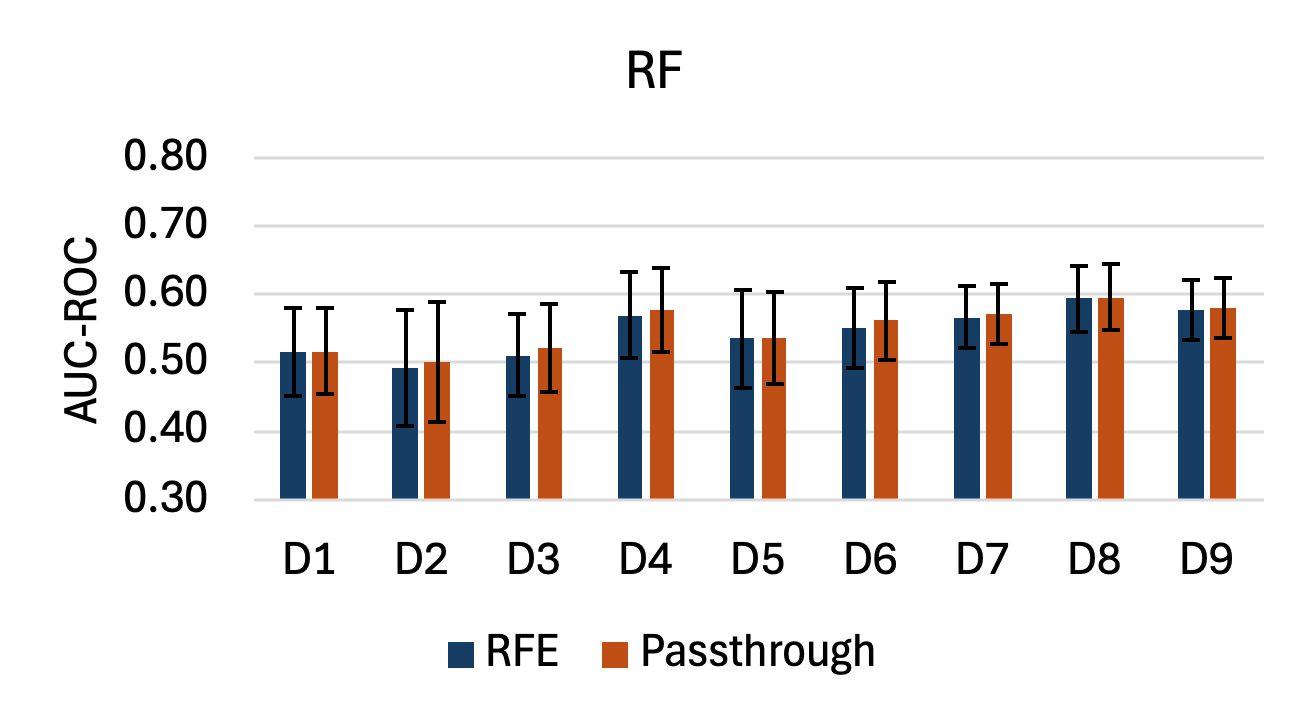 | 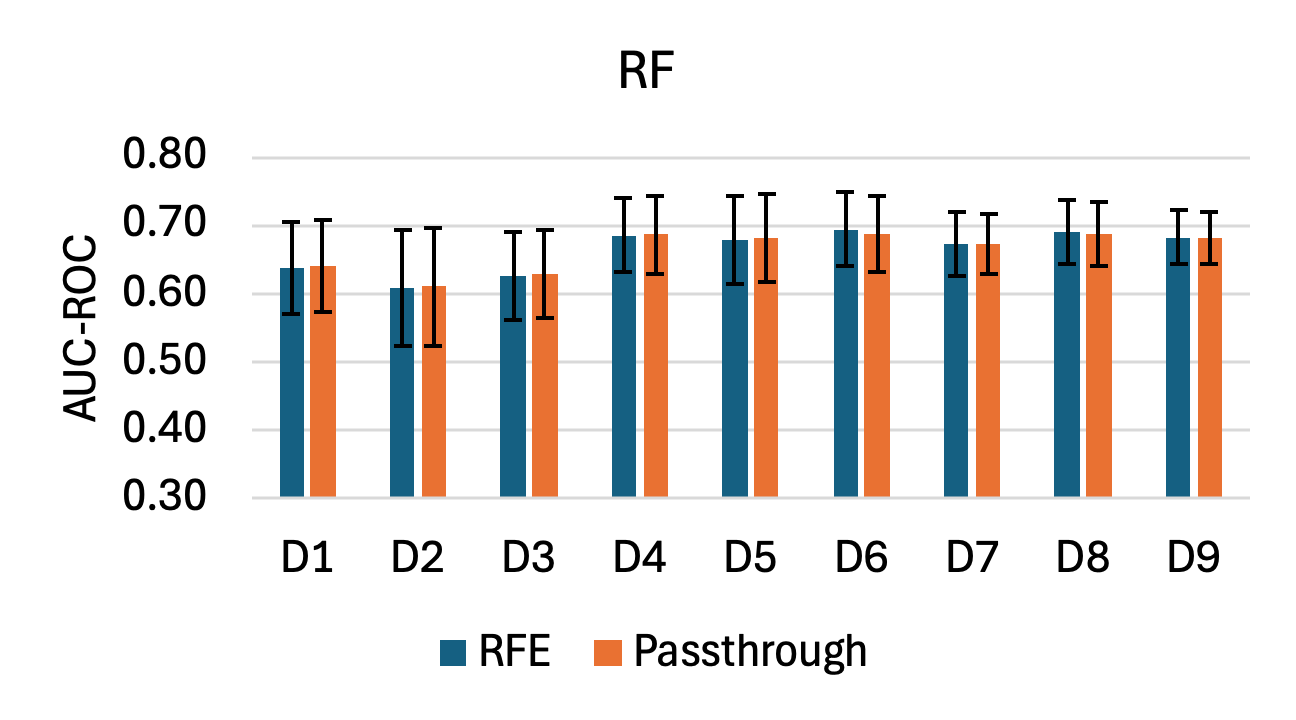 |
| 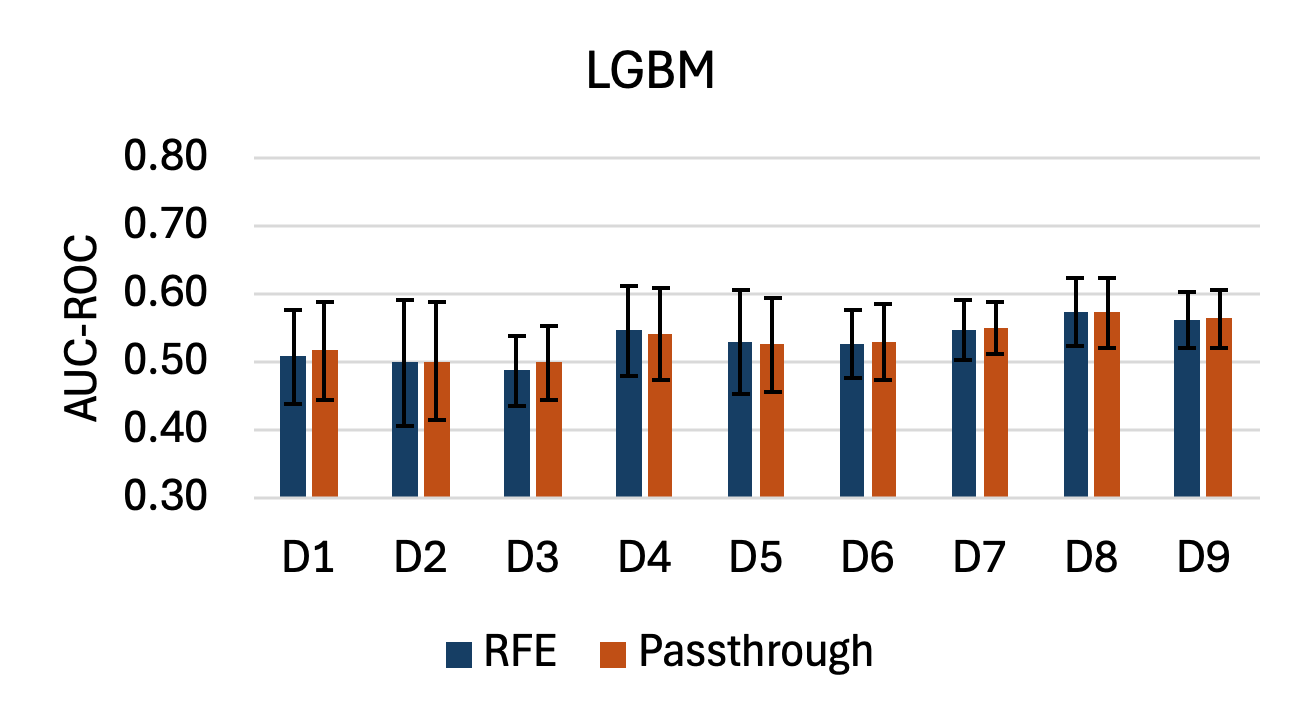 | 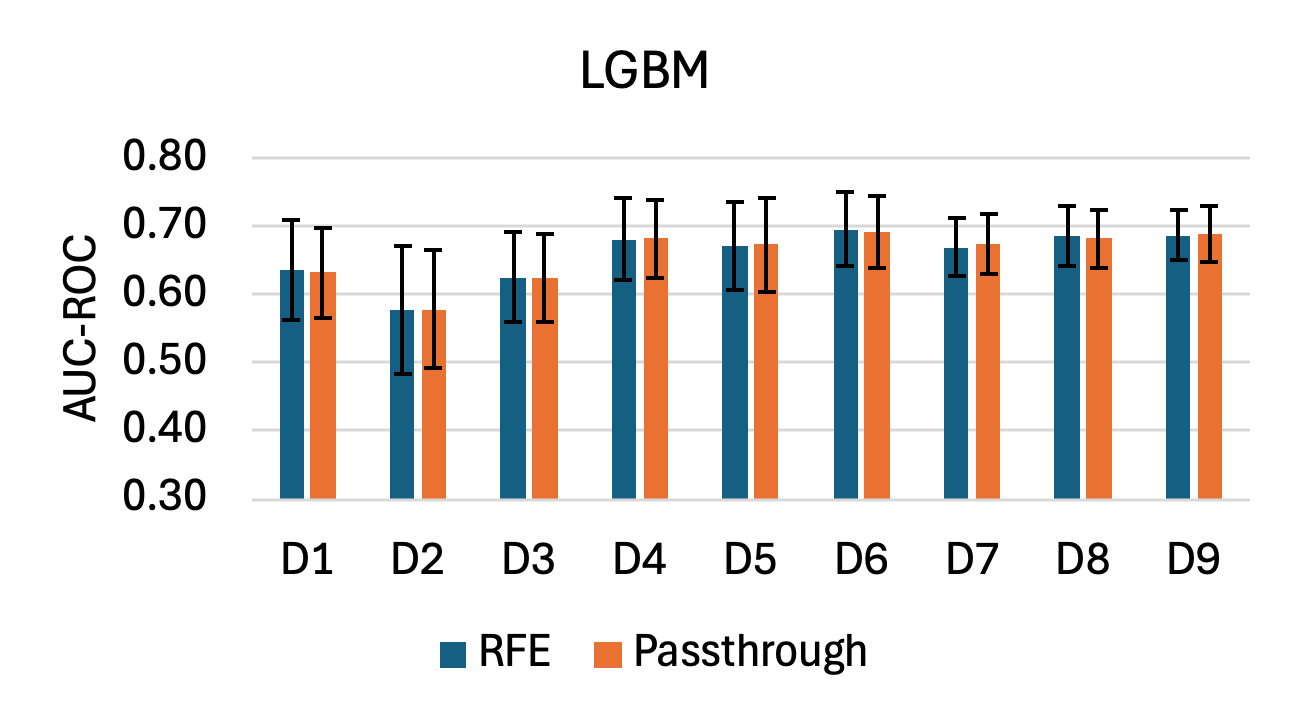 | 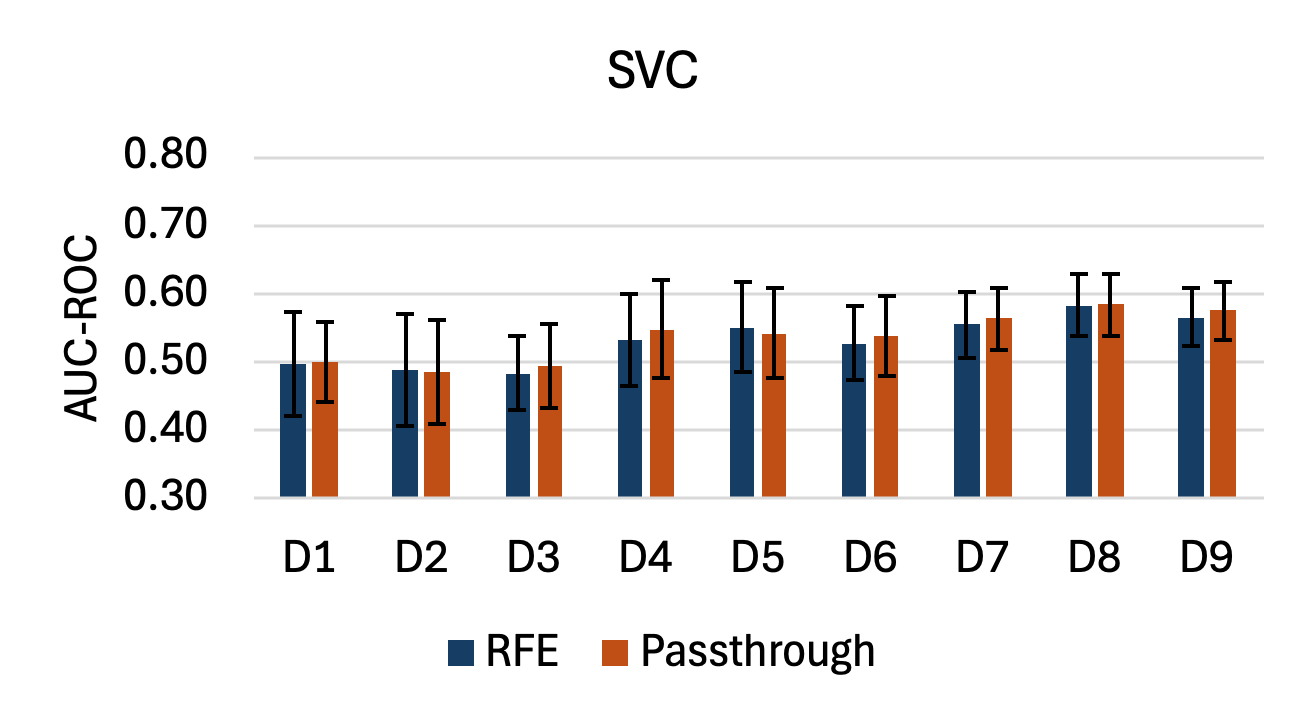 | 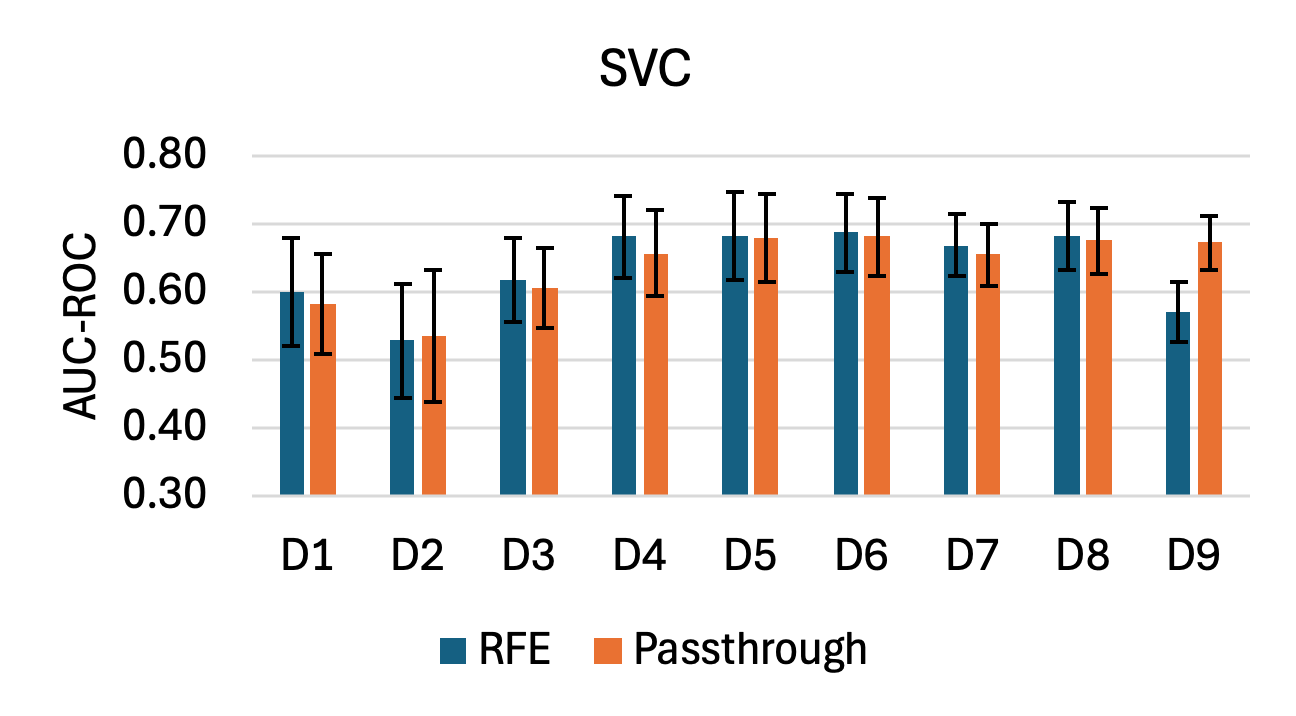 |
| 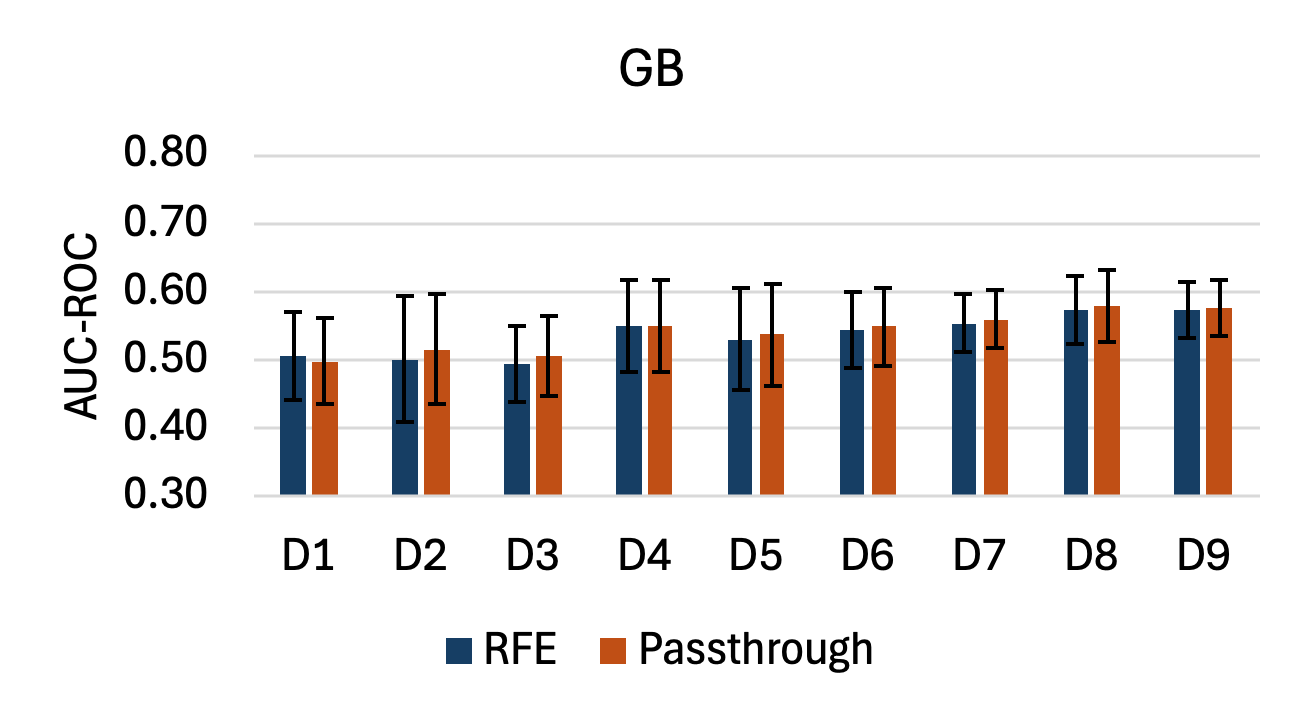 | 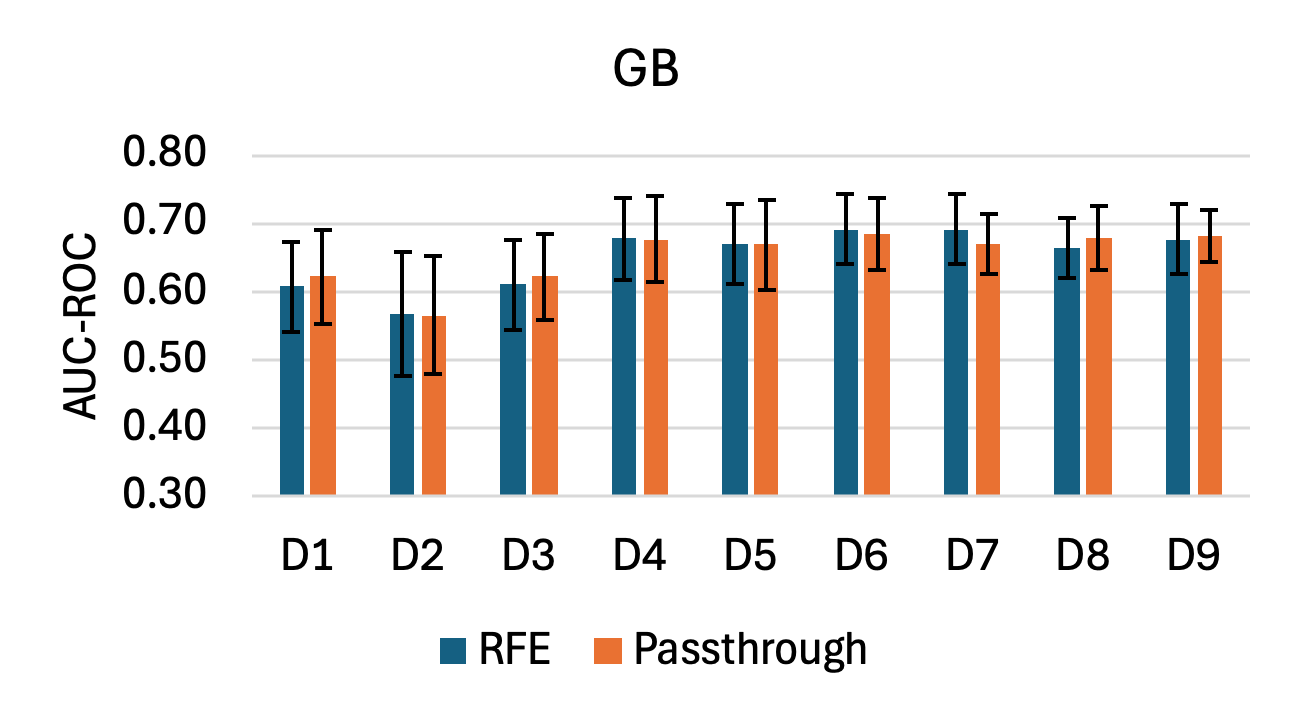 | 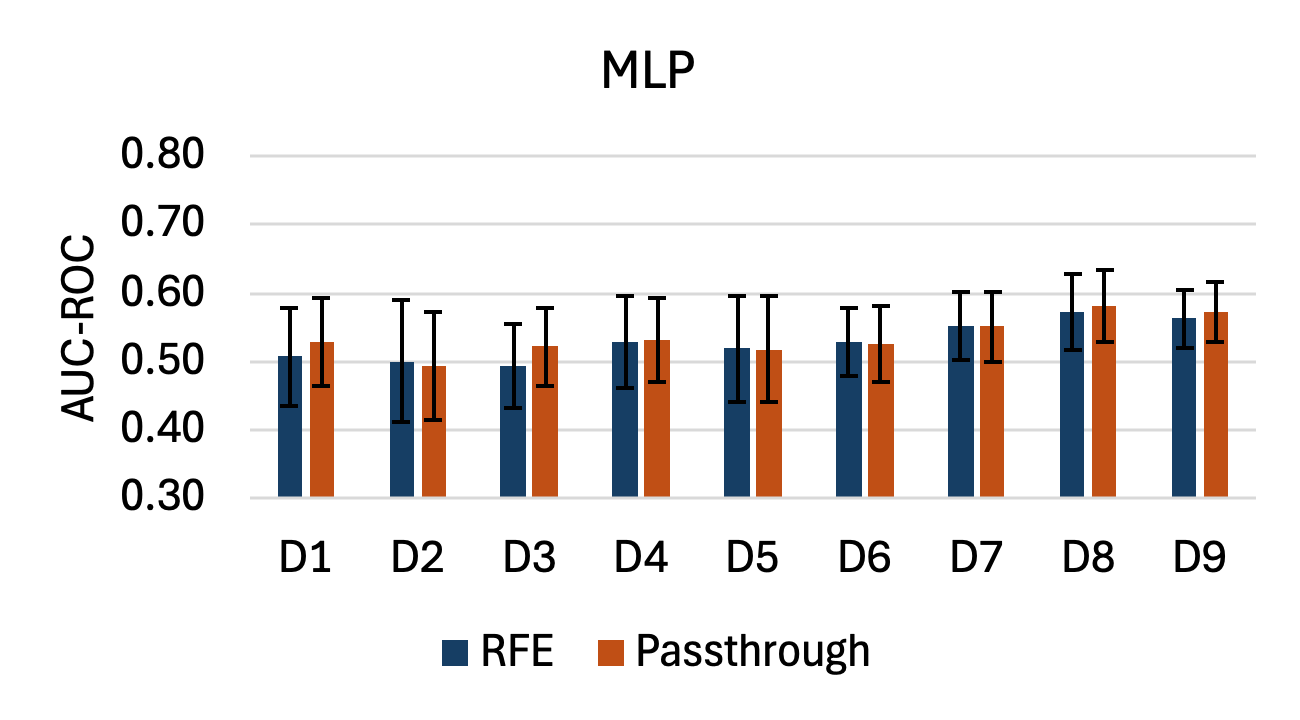 | 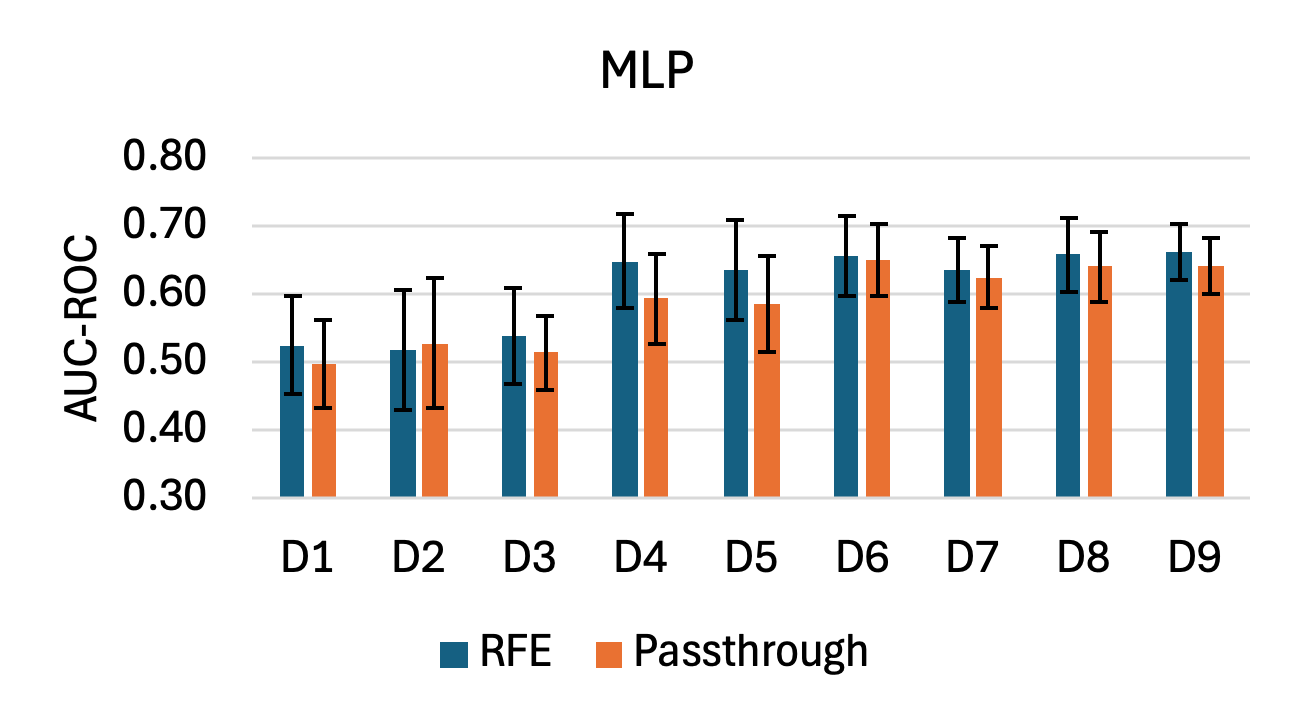 |
| 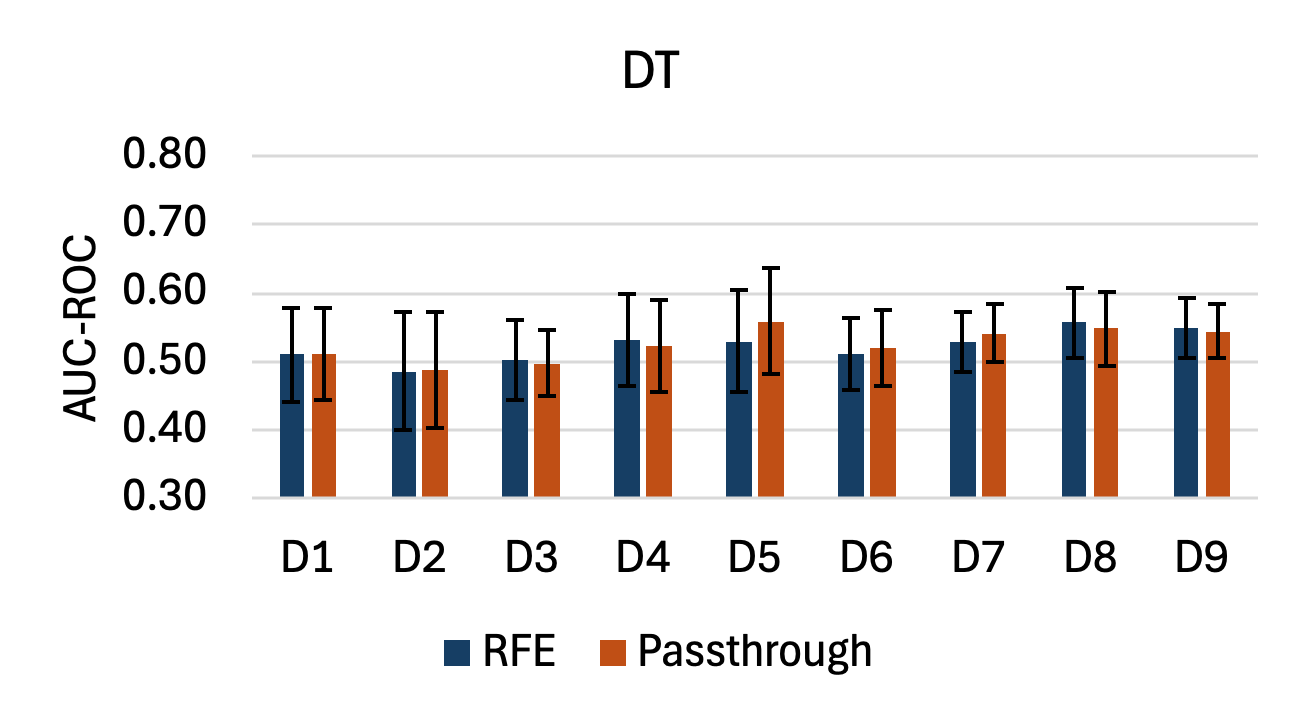 | 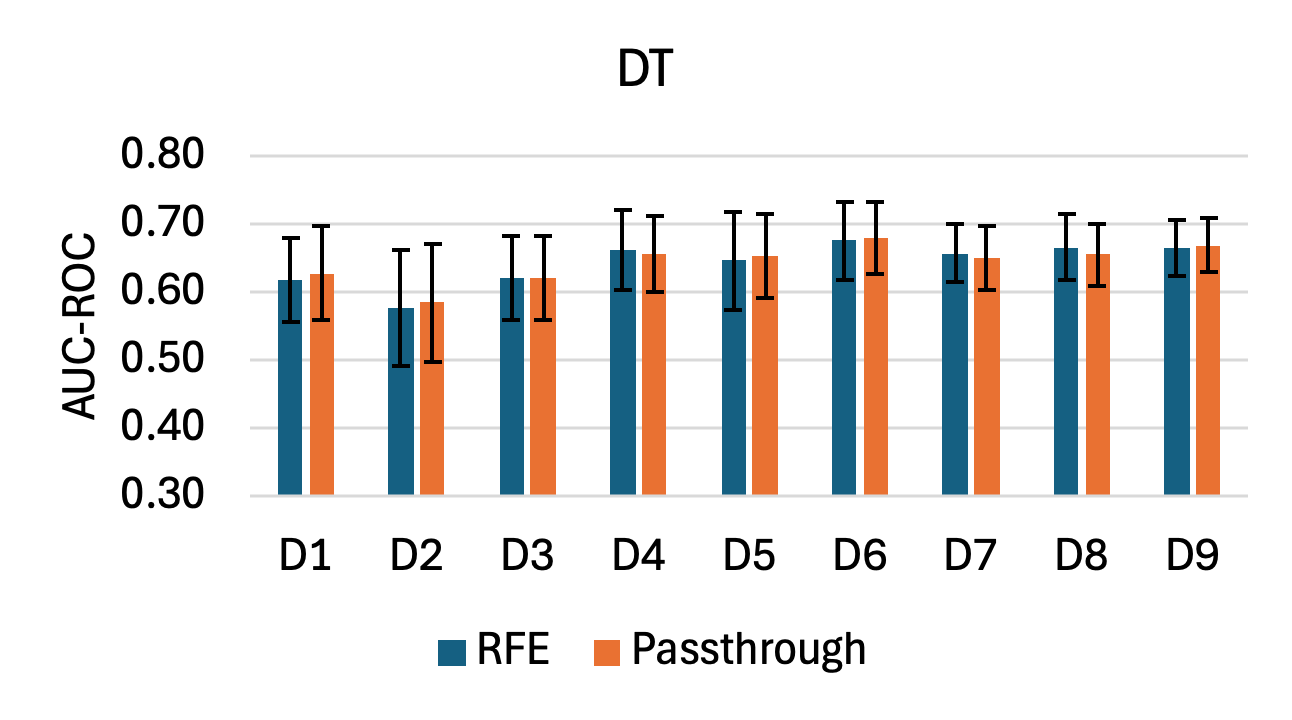 | 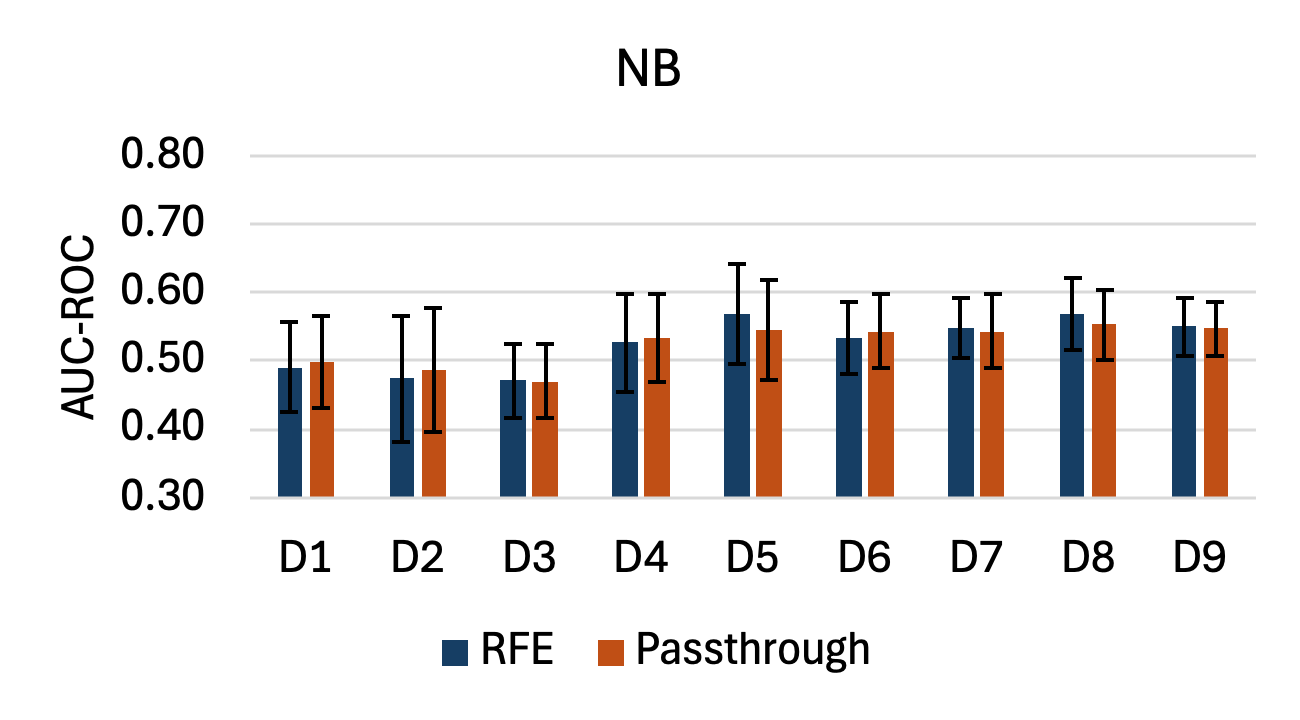 | 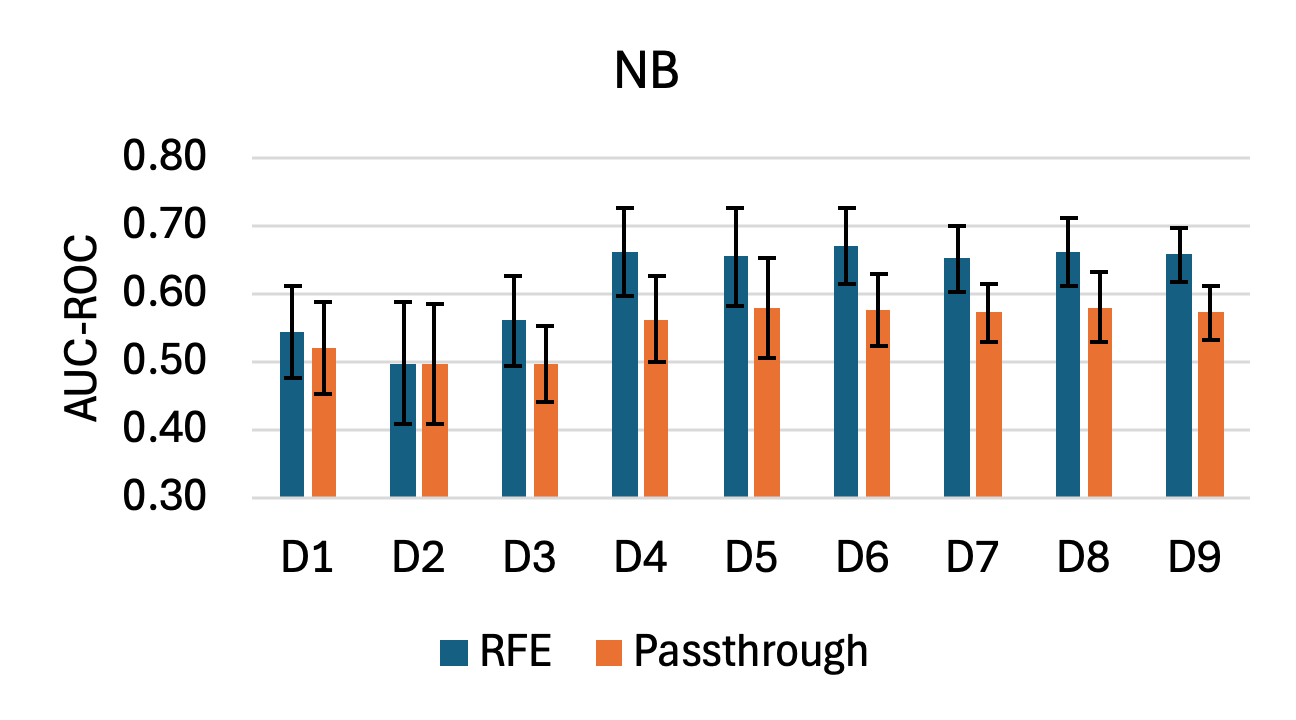 |
|  | 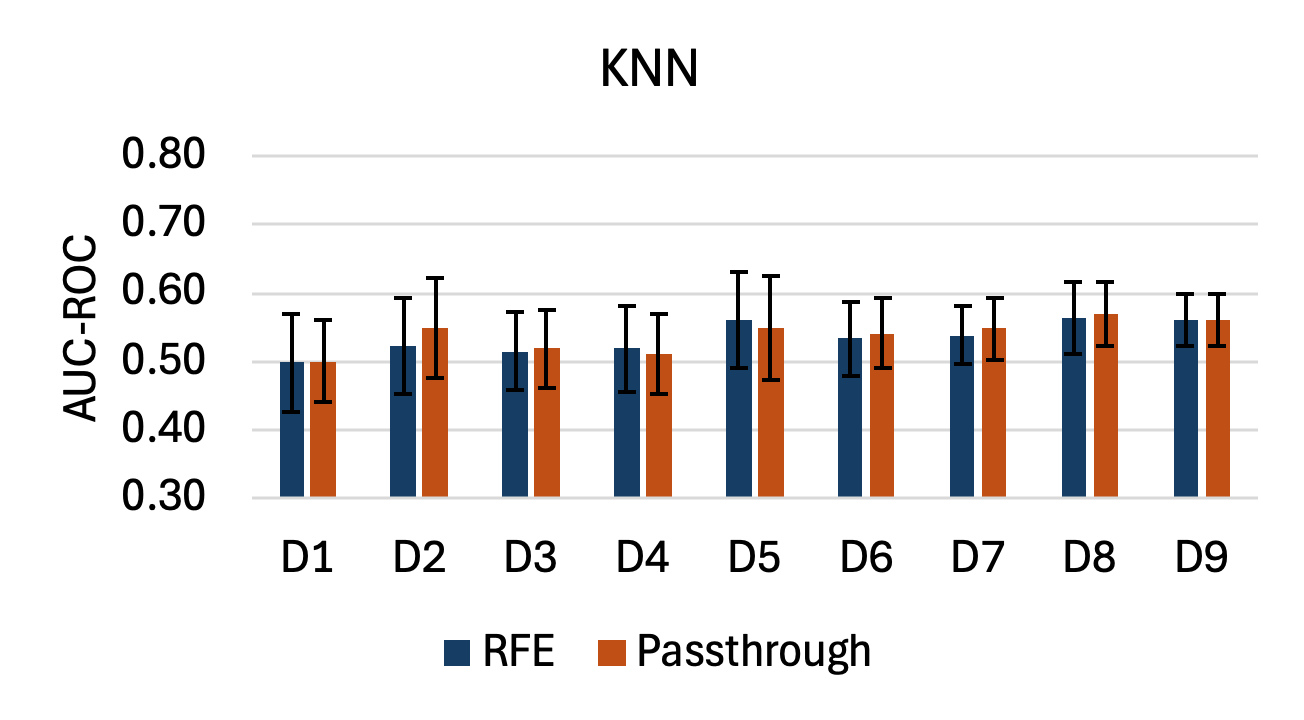 | 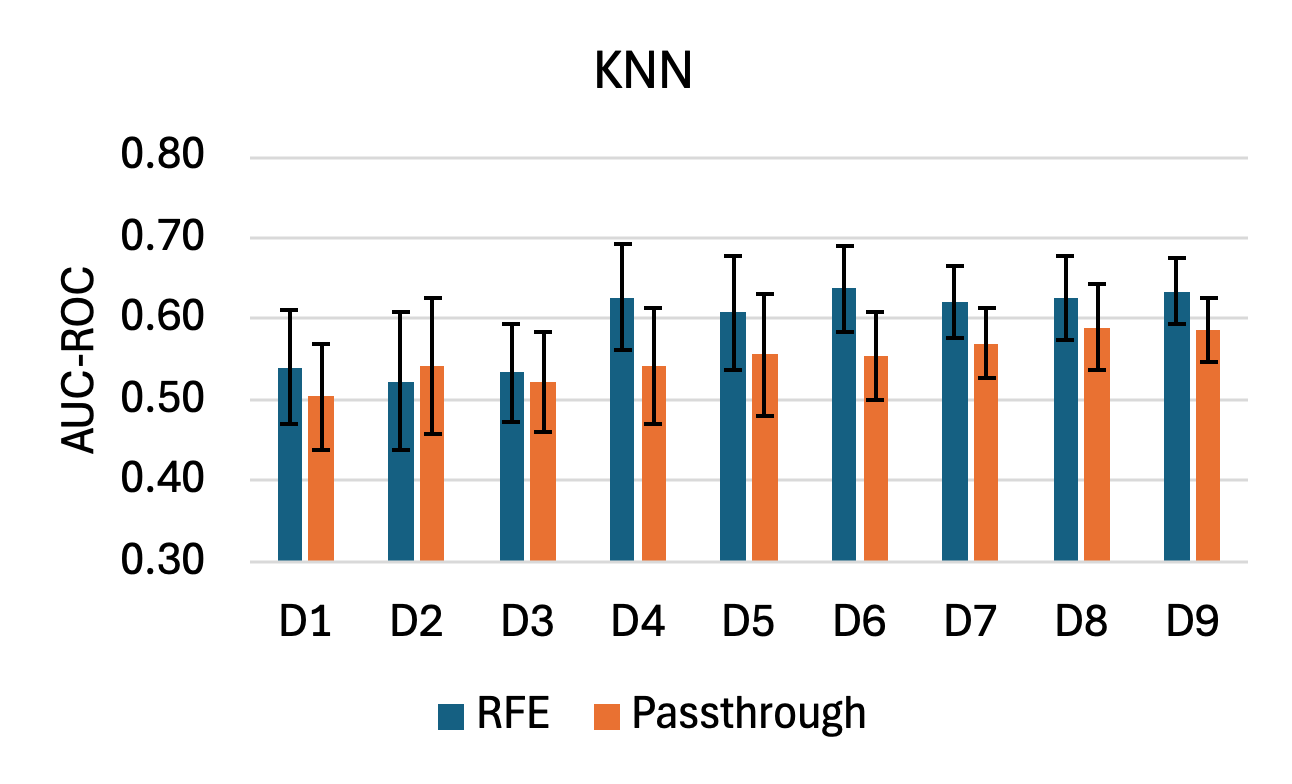 |  |

Fig. S4. AUC-ROC by classifier and modeling scenario using acoustic-only and context-augmented features, with RFE or without feature selection (Passthrough). Bars and error bars show mean $\pm$ SD across repeated outer folds.

Table S4. AUC-ROC by modeling scenario for considered feature sets without RFE (mean $\pm$ SD across classifiers).

| **Scenario** | **Acoustic-only** | **Context-augmented** |
| --- | --- | --- |
| D1 | 0.509 $\pm$ 0.011 | 0.584 $\pm$ 0.059 |
| D2 | 0.503 $\pm$ 0.020 | 0.557 $\pm$ 0.035 |
| D3 | 0.502 $\pm$ 0.017 | 0.584 $\pm$ 0.056 |
| D4 | 0.541 $\pm$ 0.019 | 0.638 $\pm$ 0.057 |
| D5 | 0.539 $\pm$ 0.012 | 0.638 $\pm$ 0.050 |
| D6 | 0.538 $\pm$ 0.013 | **0.655** $\boldsymbol{\pm}$ **0.053** |
| D7 | 0.554 $\pm$ 0.010 | 0.640 $\pm$ 0.042 |
| D8 | 0.573 $\pm$ 0.015 | 0.652 $\pm$ 0.041 |
| D9 | 0.566 $\pm$ 0.013 | 0.653 $\pm$ 0.045 |

Table S5. AUC-ROC by classifier for considered feature sets without RFE (mean $\pm$ SD across scenarios).

| **Model** | **Acoustic-only** | **Context-augmented** |
| --- | --- | --- |
| XGB | 0.538 $\pm$ 0.029 | **0.657** $\boldsymbol{\pm}$ **0.040** |
| LGBM | 0.534 $\pm$ 0.026 | **0.659** $\boldsymbol{\pm}$ **0.038** |
| GB | 0.541 $\pm$ 0.029 | 0.652 $\pm$ 0.041 |
| DT | 0.525 $\pm$ 0.024 | 0.645 $\pm$ 0.029 |
| RF | 0.551 $\pm$ 0.033 | **0.664** $\boldsymbol{\pm}$ **0.029** |
| SVC | 0.536 $\pm$ 0.036 | 0.638 $\pm$ 0.052 |
| MLP | 0.536 $\pm$ 0.028 | 0.587 $\pm$ 0.059 |
| NB | 0.525 $\pm$ 0.031 | 0.550 $\pm$ 0.035 |
| KNN | 0.538 $\pm$ 0.023 | 0.551 $\pm$ 0.028 |

| 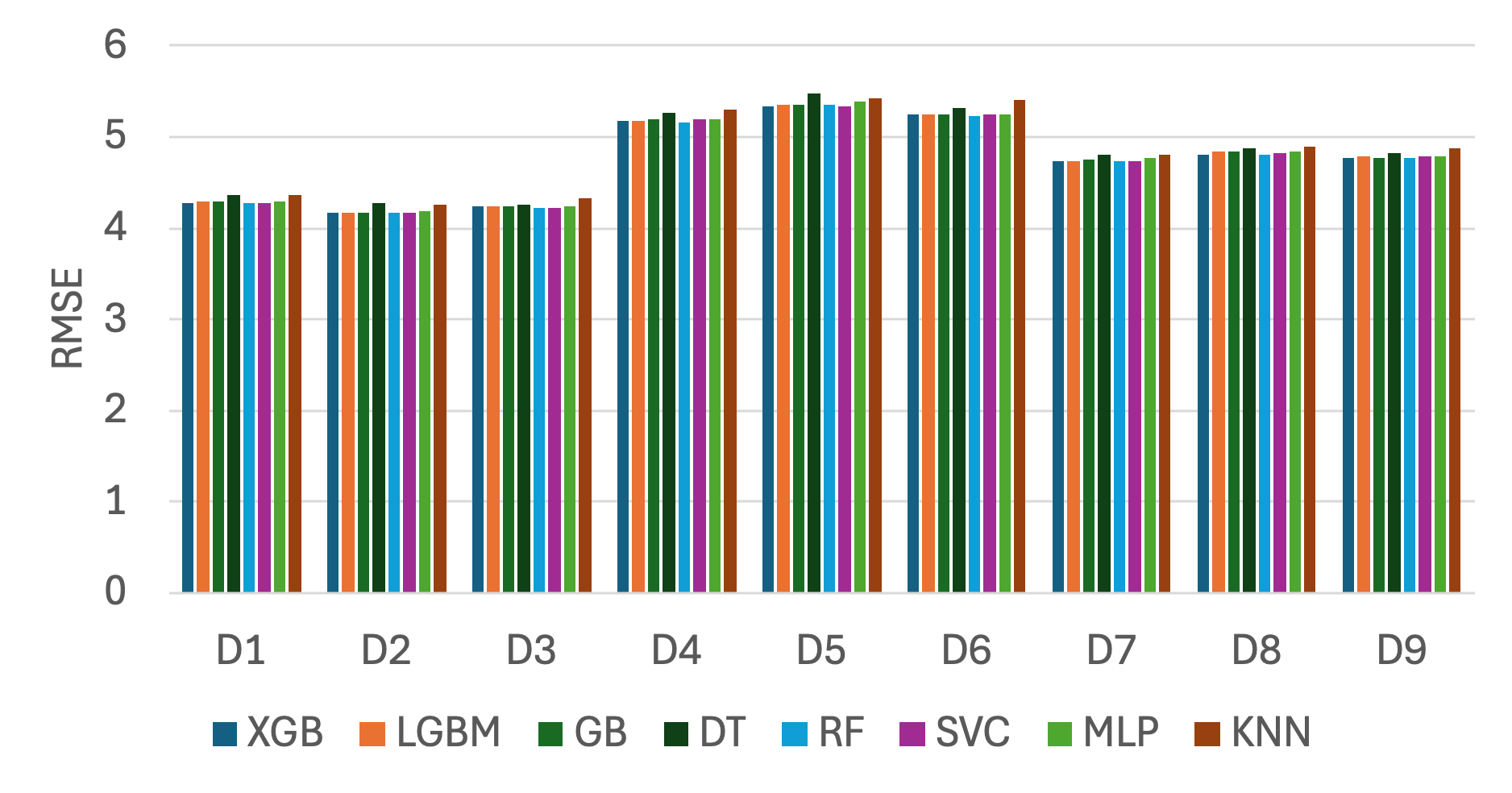  (a) | 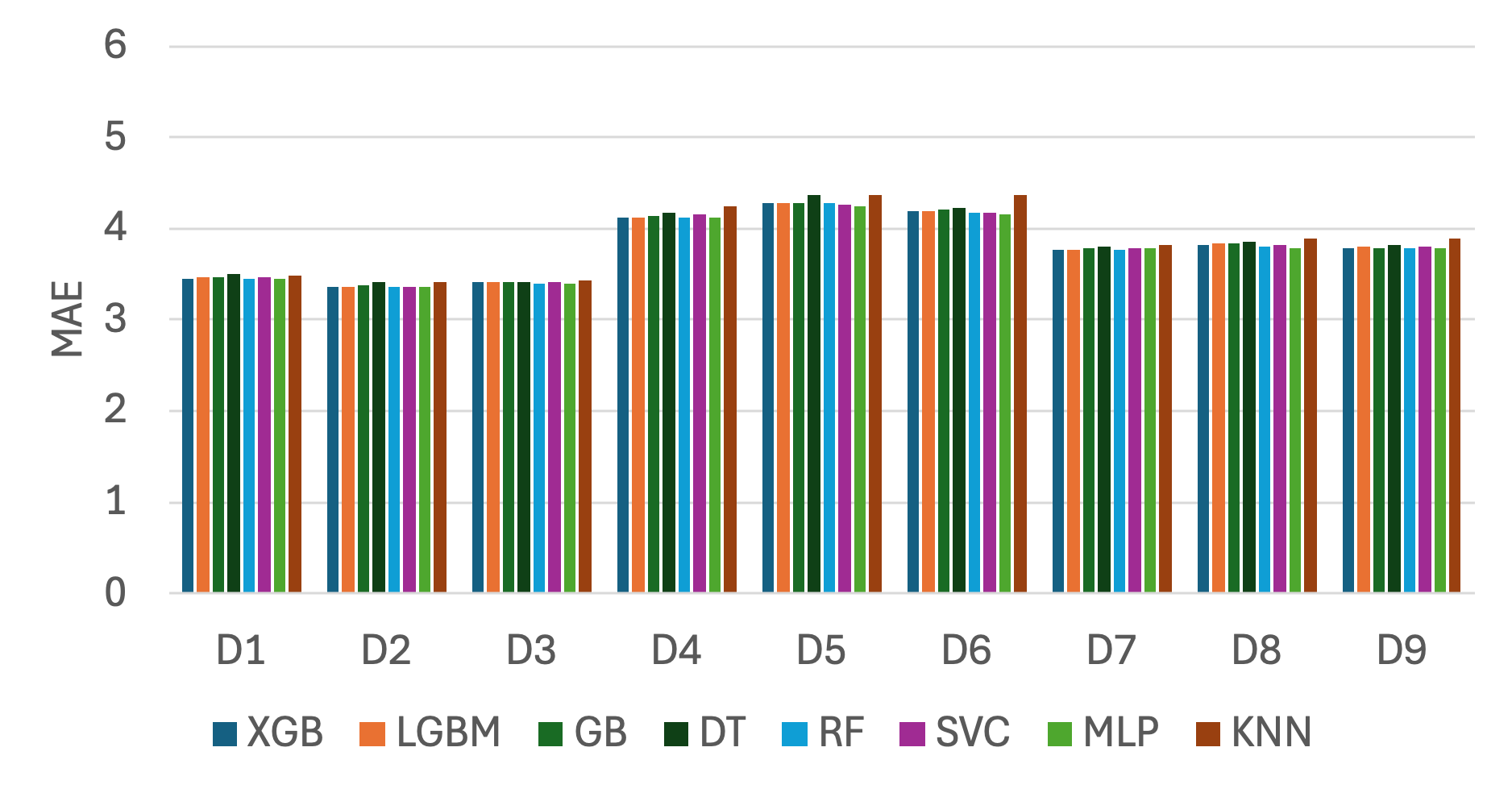  (b) |
| --- | --- |
| 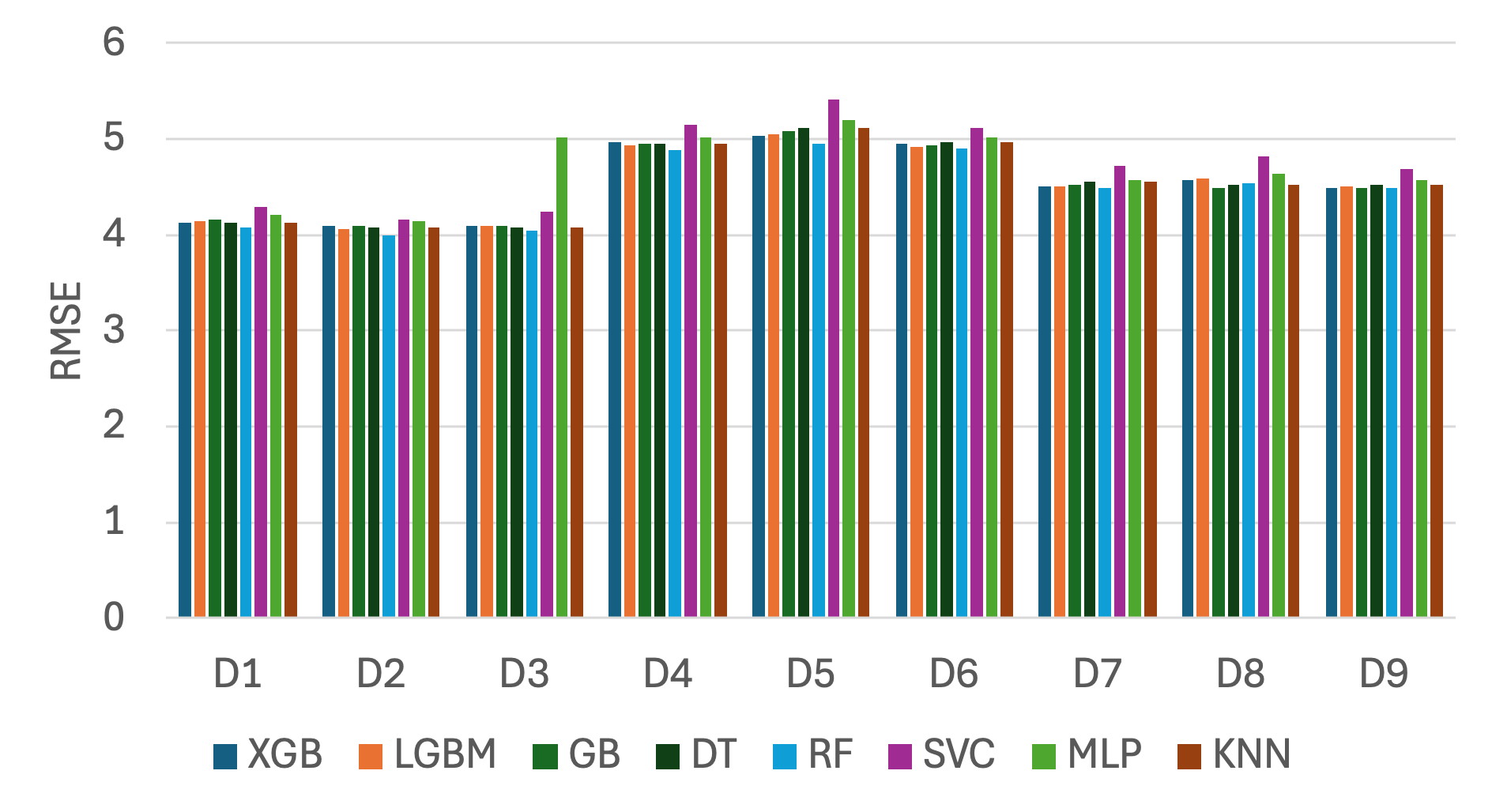  (c) | 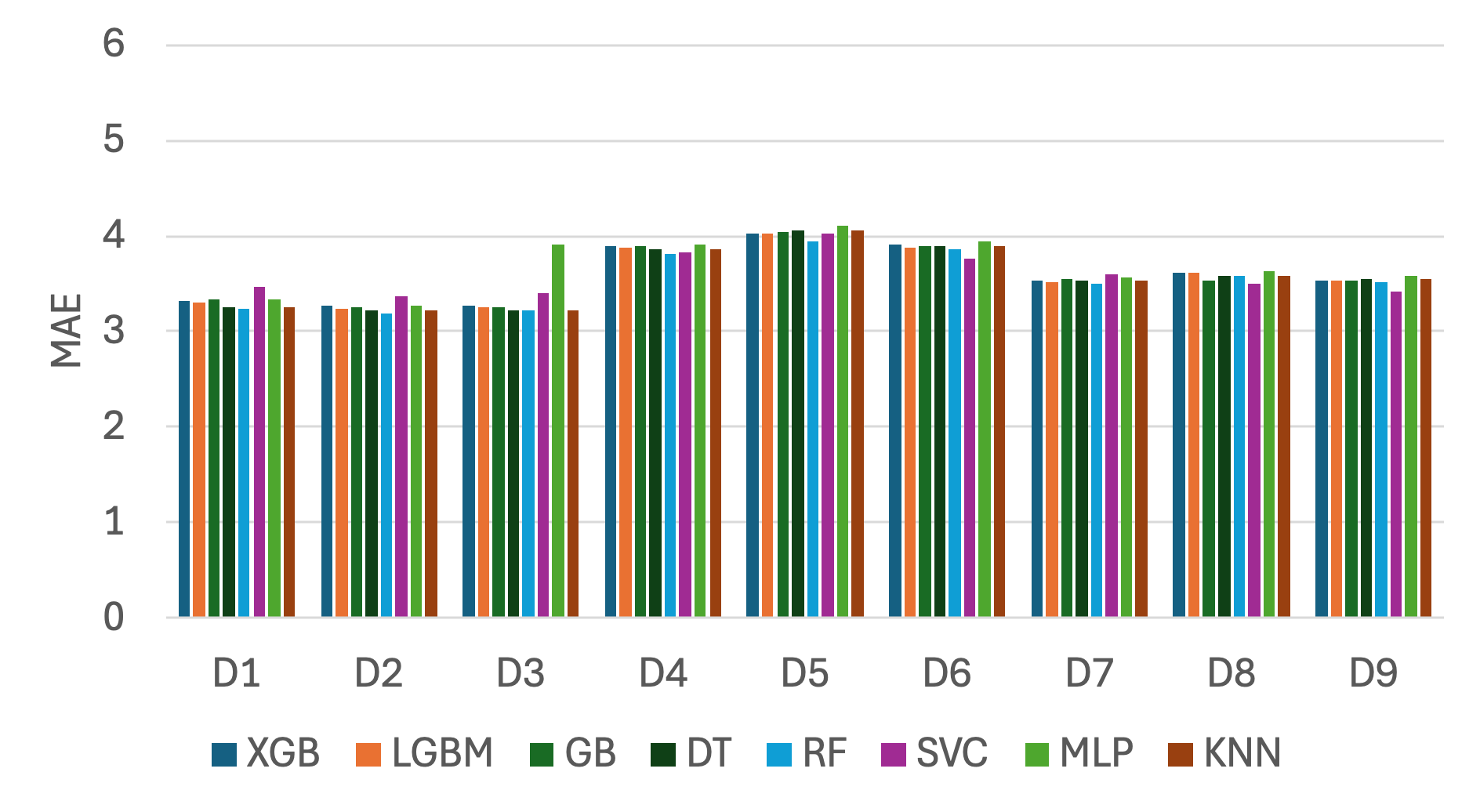  (d) |

Fig. S5. RMSE and MAE across repeated outer folds by modeling scenario for each ML model, using (a), (b) acoustic-only and (c), (d) context-augmented feature sets with RFE.

| 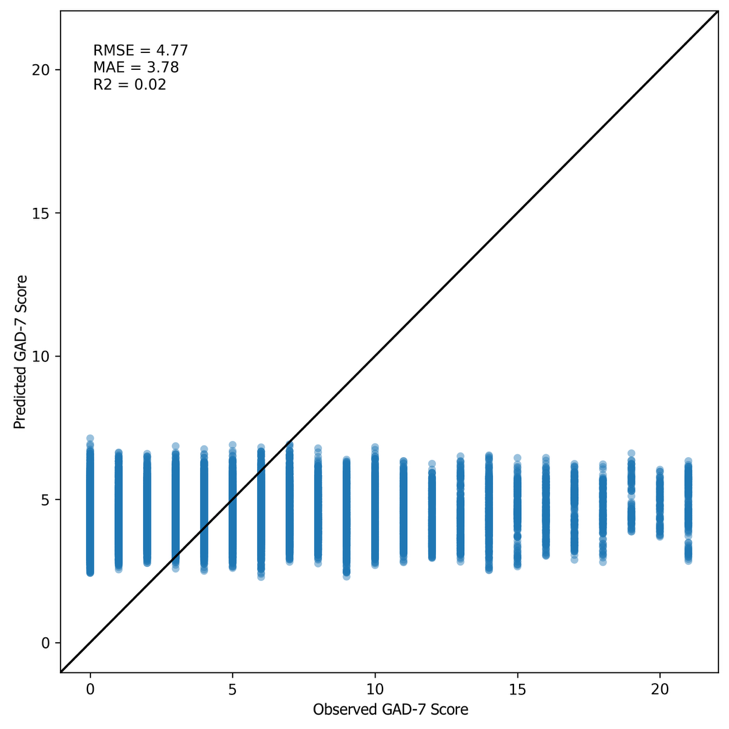  (a) | 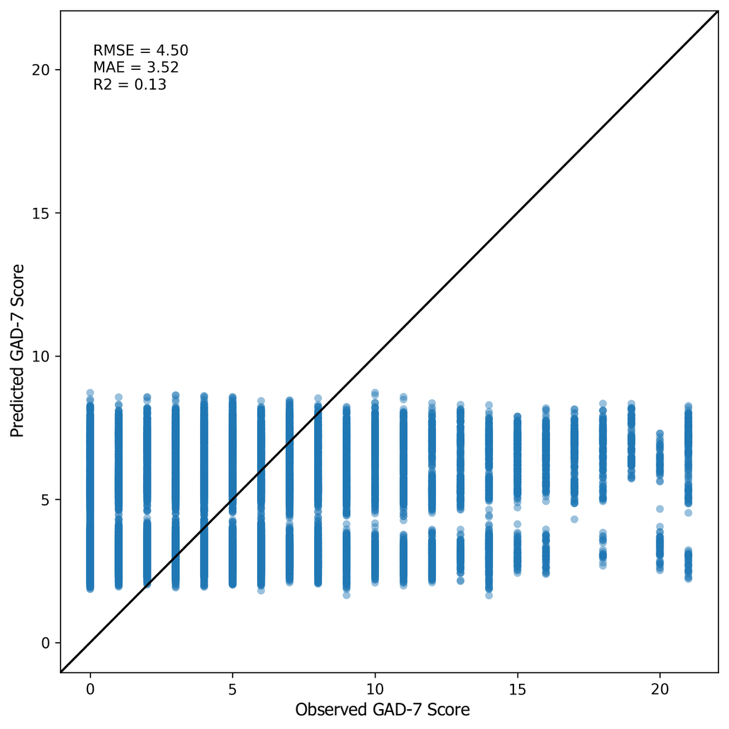  (b) |
| --- | --- |

Fig. S6. Predicted GAD-7 scores by the RF regressor in scenario D_9_ using (a) acoustic-only and (b) context-augmented feature sets with RFE.
